# Decoding mitochondrial genes in pediatric AML and development of a novel prognostic mitochondrial gene signature

**DOI:** 10.1101/2022.04.01.22273235

**Authors:** Shilpi Chaudhary, Shuvadeep Ganguly, Jayanth Kumar Palanichamy, Archna Singh, Dibyabhaba Pradhan, Radhika Bakhshi, Anita Chopra, Sameer Bakhshi

## Abstract

**Background:** Gene expression profile of mitochondrial-related genes is not well deciphered in pediatric acute myeloid leukaemia (AML). We aimed to identify mitochondria-related differentially expressed genes (DEGs) in pediatric AML with their prognostic significance.

**Methods:** Children with *de novo* AML were included prospectively between July 2016-December 2019. Transcriptomic profiling was done for a subset of samples, stratified by mtDNA copy number. Top mitochondria-related DEGs were identified and validated by real-time PCR. A prognostic gene signature risk score was formulated using DEGs independently predictive of overall survival (OS) in multivariable analysis. Predictive ability of the risk score was estimated along with external validation in The Tumor Genome Atlas (TCGA) AML dataset.

**Results:** In 143 children with AML, twenty mitochondria-related DEGs were selected for validation, of which 16 were found to be significantly dysregulated. Upregulation of *SDHC* (p<0.001), *CLIC1* (p*=*0.013) and downregulation of *SLC25A29* (p<0.001) were independently predictive of inferior OS, and included for developing prognostic risk score. The risk score model was independently predictive of survival over and above ELN risk categorization (Harrell’s c-index: 0.675). High-risk patients (risk score above median) had significantly inferior OS (p<0.001) and event free survival (p<0.001); they were associated with poor-risk cytogenetics (p=0.021), ELN intermediate/poor risk group (p=0.016), absence of RUNX1-RUNX1T1 (p=0.027), and not attaining remission (p=0.016). On external validation, the risk score also predicted OS (p=0.019) in TCGA dataset.

**Conclusion:** We identified and validated mitochondria-related DEGs with prognostic impact in pediatric AML and also developed a novel 3-gene based externally validated gene signature predictive of survival.

## Introduction

Despite recent advancements, the survival in pediatric acute myeloid leukaemia (AML) continues to remain dismal(1). Various molecular and genetic alterations are frequently used for risk stratification, identification of therapeutic targets as well as predicting disease prognosis in AML(2). Whole genome and transcriptome sequencing have been extensively used in AML to identify potential novel molecular targets and developing prognostic gene signatures to predict survival, relapse and risk stratification(3–5). However, data on potential mitochondrial genes with impact on AML are limited.

Dysregulation of mitochondrial pathways have been implicated in pathogenesis and progression of various malignancies(6). Multiple studies have reported the role of mitochondrial DNA (mtDNA) mutations, metabolic pathways and oxidative phosphorylation, on disease biology and prognosis of AML(7, 8). We have previously reported the relationship of mutations in mtDNA regulatory region with mitochondrial gene expression, and their impact on survival in children with AML(9–11). Considering the impact of mitochondrial pathways in outcome of AML, it is important to explore tumor cell heterogeneity in AML with respect to mitochondrial transcriptome and identify potential therapeutic or prognostic molecular targets.

Recently, we have reported that high mtDNA copy number is associated with poor outcome in paediatric AML and also identified its potential regulation through *PGC1A*(12). In the current study, among children with AML stratified according to mtDNA copy number, we identified mitochondria-related differentially expressed genes (DEGs) through whole transcriptome sequencing. We further validated the topmost identified mitochondria-related DEGs in a cohort of paediatric AML patients and formulated a prognostic mitochondrial gene signature for predicting survival outcome. We then validated this gene signature in an external cohort of adult AML patients from The Cancer Genome Atlas (TCGA) dataset along with estimation of predictive ability of the developed prognostic gene signature.

## Methodology

### Study design, patient population, treatment and clinical follow up

This was a prospective observational cohort study that included consecutive *de novo* paediatric (≤18 years) patients with AML registered from July 2016 to December 2019 at medical oncology outpatient clinic of our cancer centre. The workflow of the study is depicted in figure 1. Study was ethically approved by institute ethics committee (IEC/NP-336/2012, IECPG-79/22.03.2017) and informed consent was taken from care givers and assent was obtained from all participants (≥8 years). Patients with granulocytic sarcoma without marrow involvement, acute promyelocytic leukaemia, and mixed phenotypic acute leukaemia were excluded. Fifty age-matched patients of solid malignancies without marrow involvement were also enrolled as controls. Remission status and survival outcomes were noted.

**Figure 1.**
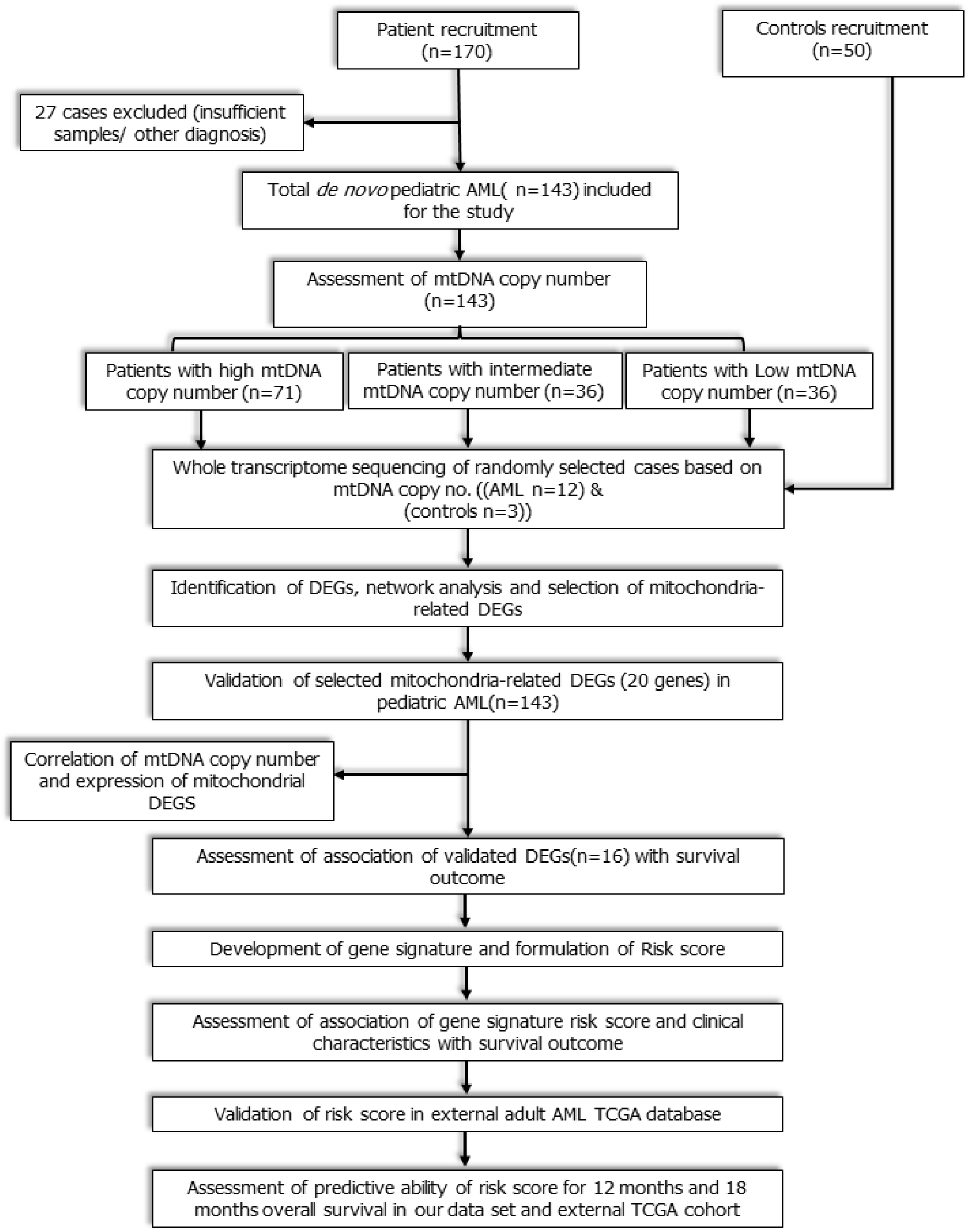
**Workflow of the study:** Study workflow showing flow of patients from enrolment to RNA sequencing, identification of mitochondria-related DEGs, development and validation of novel 3-gene based Risk score.

### Risk Assessment by karyotyping and mutation analysis

Conventional cytogenetics were done at baseline to identify translocations, inversions, deletion as well as other chromosomal abnormalities for risk stratification of pediatric AML. Mutation profiling of *RUNX1-RUNX1T1* (Runt-related transcription factor 1-RUNX1 partner transcriptional co-repressor 1 fusion transcript), *CBFB-MYH11* (Core binding factor beta-myosin heavy chain 11 fusion transcript), FLT3-ITD (Fms like tyrosine kinase 3-internal tandem duplication), and *NPM1* (Nucleophosmin 1) by reverse transcriptase polymerase chain reaction (PCR) were performed at baseline for risk assessment as per European LeukemiaNet (ELN) recommendation (13).

### Treatment protocol

All patients were treated with uniform induction protocol, i.e., 3+7 regimen including daunorubicin 60 mg/m^2^ day 1-3 and cytarabine 100mg/m^2^ continuous infusion day 1-7. Consolidation therapy with three cycles of high dose cytarabine at 18g/m^2^ were given to patients after achieving complete remission (CR) whereas repeated induction with ADE regimen (cytarabine: 100 mg/m2 twice daily, day 1-10; daunorubicin: 50 mg/m2, day 1-3; and etoposide: 100 mg/m2, day 1-5) were used for refractory or relapse cases. Patients at CR2 underwent allogeneic hematopoietic stem cell transplantation with matched sibling donor if available (13, 14).

### Isolation of bone marrow mononuclear cells (BMMCs), DNA and RNA extraction and quantification

BMMCs were isolated by Ficoll-Hypaque (Sigma diagnostics, USA) density gradient centrifugation followed by isolation of total RNA and DNA. Briefly, BM samples collected from all the patients and controls were layered onto the histopaque in 15 ml falcon tube followed by centrifugation at 400g for 30 min at room temperature. The mononuclear cells layer was carefully taken out after centrifugation and washed twice with phosphate buffer saline (PBS). BMMC were then stored for DNA isolation using isopropanol precipitation method and total RNA was isolated using TRIzol method as per manufacturer’s protocols. Quality of DNA and RNA was checked by Nanodrop 1000 (Thermo Fisher) and integrity of RNA was checked by TapeStation (Agilent).

### Estimation of mitochondrial DNA copy number

Relative mtDNA copy number was assessed by fluorescent DNA binding dye SYBR based quantitative real time PCR using, with Roche Light Cycler 480 II. The relative mitochondrial DNA copy number was normalized to expression of nuclear gene *ACTB*, which was chosen as a nuclear housekeeping gene. The mitochondrial DNA copy number, normalized to copies per cell, in each subject and control sample was calculated using the following formula: 2^[Ct(*β*−actin) −Ct (minor arc)] (Ct being respective cycle thresholds). Relative mitochondrial DNA copy numbers of patients were then compared with controls.

### Whole transcriptome sequencing, identification of differentially expressed genes (DEG) and selection of DEG for analysis

All the samples were classified into three separate groups based on relative mtDNA copy number(12). Patients were categorized into: AMLCN_H (mtDNA copy number ≥ 75^th^ percentile), AMLCN_I (mtDNA copy number 50th to 75th percentile) and AMLCN_L (mtDNA copy number < 50th percentile) groups. A subset of samples was randomly selected from each of the three sub-groups and controls with RNA integrity score above 7 and a total of 15 samples (12 patients including 3 from AMLCN_H group, 4 from AMLCN_I group, 5 from AMLCN_L group and 3 controls) were sent for whole transcriptome profiling for the identification of DEGs compared to controls.

### Library Construction and sequencing

The sequencing library was prepared by random fragmentation of the cDNA sample, followed by 5’ and 3’ adapter ligation, after end-repair and the addition of an ‘A’ base and SPRI clean-up. The prepared cDNA library was amplified using PCR for the enrichment of the adapter-ligated fragments. The individual libraries were quantified using a NanoDrop spectrophotometer (Thermo Scientific) and validated for quality with a Bioanalyzer (Agilent Technologies). Adapter-ligated fragments were then PCR amplified and gel purified. cDNA library was used for sequencing which was carried out on Illumina Hiseq 4000 NGS platform.

### Processing of the reads and differential expression analysis

The quality of raw reads was first checked by FastQC version v0.11.8. Trimmomatic was performed to remove adapter sequences and low-quality reads for further analysis. The trimmed reads were then aligned to reference Human genome (hg38) using HISAT2 tool. SAM tool was used to convert SAM files into BAM files which was used for quantification and estimation of aligned reads by StringTie (v2.0.6). Lastly differential expression analysis was performed by limma Bioconductor package (version 3.48.1). Absolute fold change value ≥ 2 (a ≥ two-fold change in expression, either upregulated or downregulated) and adjusted p value (q ≤ 0.05) threshold was used for the identification of differentially expressed genes (DEGs). Volcano plots were made with log fold change; p value and adjusted p value of the transcripts were calculated using devtools in R package (version 3.6.1). Gene name of the corresponding transcripts with significant p value, adjusted p value and log 2-fold difference compared to controls, were fetched using gProfilerR library in R package. Exclusive differentially expressed genes were identified among the groups by making Venn diagram using Venny (Venny 2.1) online tool. (Figure 2).

**Figure 2:**
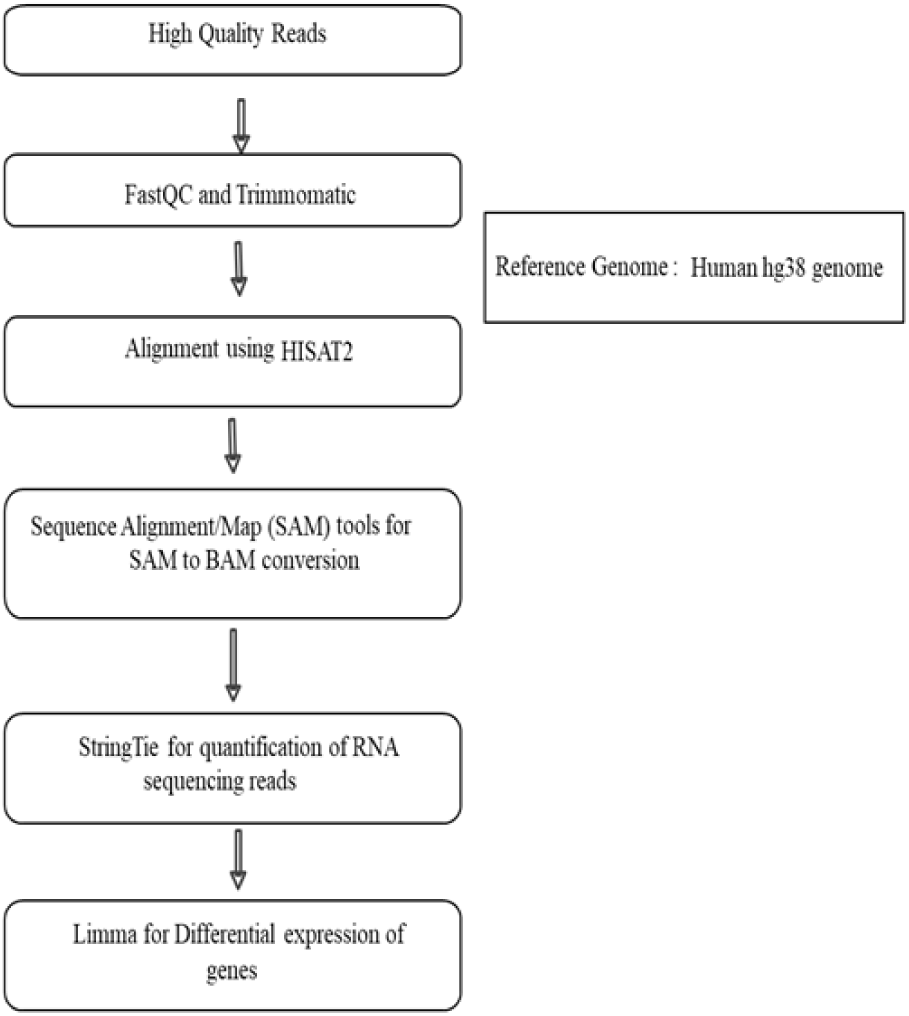
Workflow of RNA sequencing data analysis using various bioinformatic tools

### Construction of protein-protein interaction network, determination of HUB genes and MCODE analysis

A interactive network of proteins encoded DEGs present in the respective groups were constructed using Search Tool for the Retrieval of Interacting Genes/Proteins (STRING) database (15) in Cytoscape. Cytohubba plugin was used for identifying Hub genes among the whole interactome network. Those genes which got enriched using at least six different topological algorithms among Degree, Edge Percolated Component (EPC), Maximum Neighbourhood Component (MNC), Density of Maximum Neighbourhood Component (DMNC), Maximal Clique Centrality (MCC) and centralities based on shortest paths, such as Bottleneck (BN), Eccentricity, Closeness, Radiality, Betweenness, and Stress were considered as Hub genes(16). Cytoscape plug-in MCODE clustering algorithm was used for identifying the maximum ranked cluster which has a highly interconnected region, also known as seed nodes as well as their neighbour nodes in the whole network (17). Subcellular location of the genes was identified from the gene ontology table. Compartment mitochondria score was used for selecting mitochondrial related genes among all the AML subgroups.

### Selection and validation of mitochondria-related DEGs

Out of all identified DEGs from transcriptome sequencing, mitochondria-related genes were filtered using Cytoscape compartment mitochondrion score (0 being minimum and 5 being highest)(18). DEGs with topmost mitochondrial compartment score were selected for validation in a cohort of paediatric patients with AML. The genes of MCODE cluster 1 and Hub genes were assessed for their mitochondrial localization as above and genes in each group with highest mitochondrial compartment score were selected for validation. Based on these selection strategies, a total of 20 mitochondria-related DEGs were selected for validation. Real time PCR was performed to validate the selected mitochondria-related genes using specific primers (Table S1) and the gene expressions were quantified per previously described protocol(12). All experiments were replicated in triplicates.

### Comparison of validated mitochondria-related DEGs in TCGA data set

For external validation of mitochondrial related DEGs, the RNA-sequencing data (Illumina HiSeq 2000) of TCGA adult AML(LAML) dataset was chosen, which is one of the largest datasets of transcriptomic profile in AML with recorded clinical outcome(https://www.cbioportal.org/study/summary?id=laml_tcga). The adult dataset was specifically chosen to see the impact of prognostic impact of the validated mitochondria-related age group in a different age group as well. The expression of validated DEGs was compared with LAML data set using online available GEPIA2 (Gene Expression Profiling Interactive Analysis) web server (http://gepia2.cancer-pku.cn/#index)(19).

## Statistical Methods

### Prognostic impact of mitochondria-related DEGs and development of mitochondrial gene signature

Statistical analysis was carried out in SPSS (v23, IBM, NY, USA). Descriptive statistics were used to summarize baseline characteristics. Gene expression was reported as median values with interquartile ranges. Gene expression values and clinical continuous variables with non-parametric distribution were compared by Mann Whitney test. Clinical categorical variables were compared by Chi-square test/Fisher’s exact test as applicable. Alpha error was adjusted for multiple comparisons by Bonferroni correction. Kaplan Meier method was used to analyse time to event outcomes. Duration from enrolment to relapse or death due to any cause was considered as event free survival (EFS). Time from enrolment to death due to any cause was defined as overall survival (OS). Survival data was censored till 31^st^ Dec 2020. The follow-up estimation was done by reverse Kaplan Meier method.

Prognostic impact of all validated DEGs on OS of the whole validation cohort was performed by multivariable Cox regression analysis in a forward stepwise manner based on log likelihood change. Validated DEGs with significant (p<0.05) predictive impact on OS in multivariable analysis were included for the prognostic gene signature model. The proportional hazard assumption was assessed by Schoenfeld global test. Internal validation of the multivariable prognostic model was carried out by bootstrapping method (10000 resampling) and genes that did not satisfy bootstrapping validation were excluded. A prognostic risk score was generated using cox regression coefficient Beta (β) values of included genes, of the final multivariable model as below:

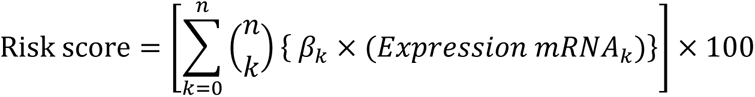

The area under the time-dependent receiver operating characteristic (ROC) curve (Timed AUC) for 12-months and 18-months survival was estimated and Harrel’s C-index of the prognostic model was calculated using the R package “survminor” in R (version 4.0.3). Patients were classified into two groups based on their risk score above (High-risk) and below (Low-risk) the median. The survival outcomes of the patients were compared between high-risk score vs low-risk score patients using Kaplan Meier analysis to evaluate the prognostic significance of the gene signature model.

### Impact of clinical features and independent prognostic value of the gene signature

The role of demographic and clinical features, including gender, age, haemoglobin, hyperleukocytosis (≥50000/µl), platelet count, presence of chloroma and ELN risk stratification(2)on survival outcome was analysed using the Cox regression. Factors with p<0.1 in univariable analysis were included for multivariable Cox regression in a forward stepwise manner using log likelihood change. Clinico-demographic factors which were significant in multivariable analysis were included in a multivariable Cox regression model along with gene signature risk score to explore the independent predictive value of gene signature. The timed AUC using 12-months survival and 18-months survival as the outcome and Harrel’s C-index of the clinical prognostic model and combined clinical and gene signature prognostic model were compared for identifying the additional prognostic benefits of gene signature over clinical parameters. The impact of mtDNA copy number on survival outcome was also analysed similarly.

### External validation of mitochondrial prognostic gene signature in TCGA dataset

The prognostic impact of our gene signature risk score on OS was done in TCGA LAML (n=179) dataset by Cox regression analysis. Patients were similarly sub-grouped into high-risk and low-risk category based on median value of the gene signature; the survival outcomes of the patients were compared between high-risk score vs low-risk score patients using Kaplan Meier analysis and timed AUC of 12-month and 18-month survival was evaluated. Based on available karyotyping data, patients of the TCGA dataset were grouped into poor-risk karyotype and others (including good and intermediate-risk karyotype). The karyotype category and mitochondrial gene risk score were assessed for their impact on OS by a multivariable Cox regression model to explore the independent predictive value of the gene signature in the external cohort as well.

## Results

### Patients’ recruitment and baseline clinical features

Total 170 patients were enrolled, out of which 27 patients (5 patients were AML M3, 4 had granulocytic sarcoma without marrow involvement, and 18 patients had insufficient samples) were excluded. The baseline demographic and clinical characteristics of final 143 patients are summarized in Table 1. Median age was 10 years (range: 0.8-18 years) and 50% of the patients were classified as ELN good risk. Total 104 patients (72.7%) achieved complete remission (CR) after induction therapy. At median follow-up of 36 months (32.67-39.33 months), the median OS was 21.93 months (13.54–30.31months). The clinical characteristics of the TCGA LAML dataset are summarized in Table S2.

**Table 1:**
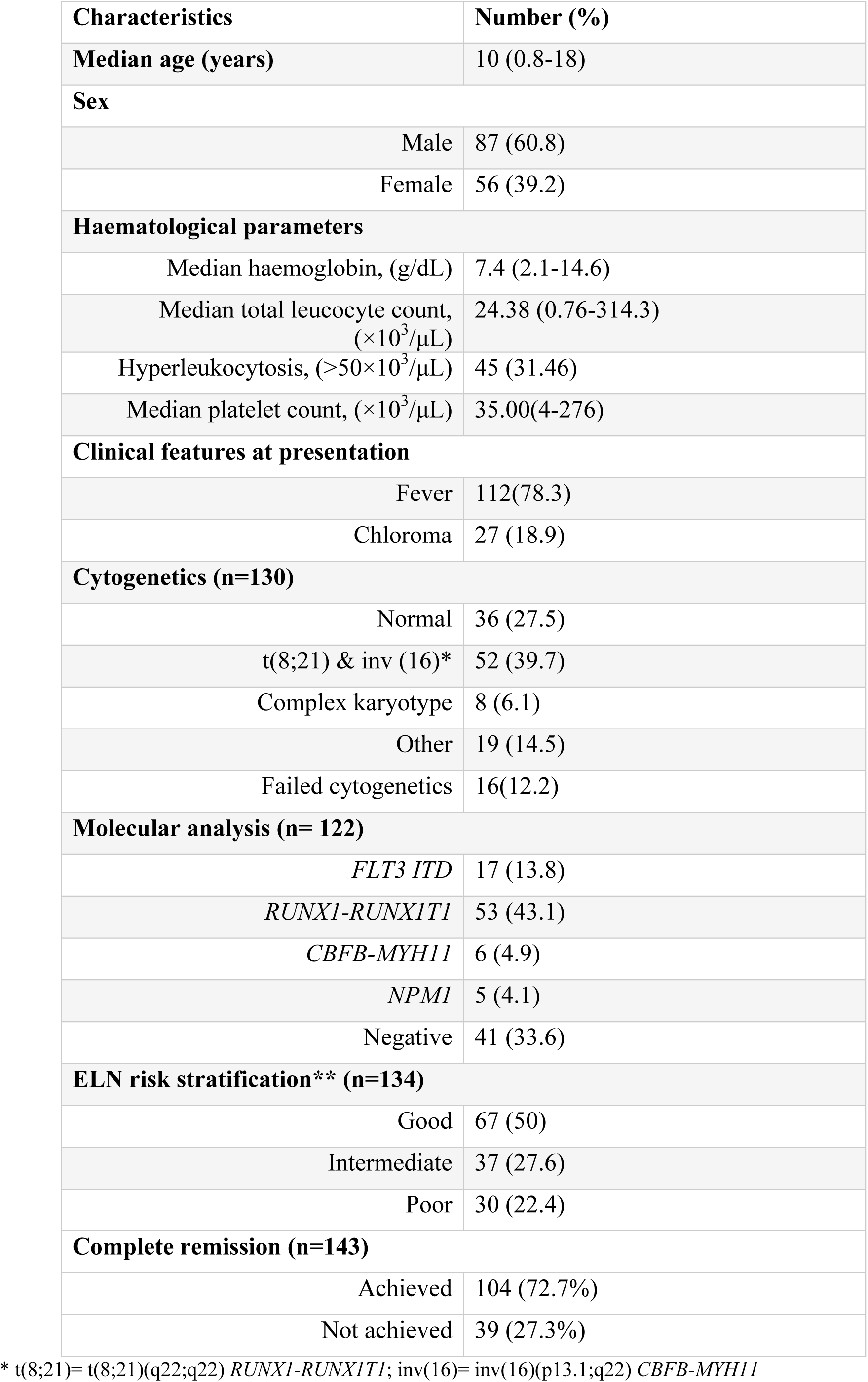
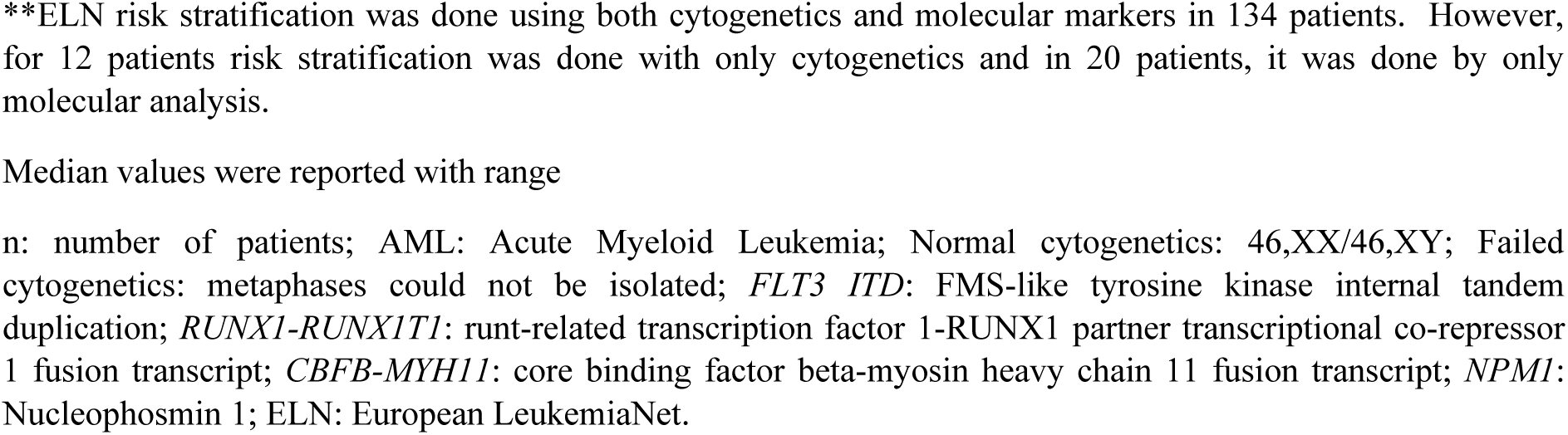
Baseline characteristic features of pediatric AML patients (n=143)

| Characteristics | Number (%) |
| --- | --- |
| Median age (years) | 10 (0.8-18) |
| **Sex** |  |
| Male | 87 (60.8) |
| Female | 56 (39.2) |
| **Haematological parameters** |  |
| Median haemoglobin, (g/dL) | 7.4 (2.1-14.6) |
| Median total leucocyte count, ($\times 10^3/\mu\text{L}$) | 24.38 (0.76-314.3) |
| Hyperleukocytosis, ($>50 \times 10^3/\mu\text{L}$) | 45 (31.46) |
| Median platelet count, ($\times 10^3/\mu\text{L}$) | 35.00(4-276) |
| **Clinical features at presentation** |  |
| Fever | 112(78.3) |
| Chloroma | 27 (18.9) |
| **Cytogenetics (n=130)** |  |
| Normal | 36 (27.5) |
| t(8;21) & inv (16)* | 52 (39.7) |
| Complex karyotype | 8 (6.1) |
| Other | 19 (14.5) |
| Failed cytogenetics | 16(12.2) |
| **Molecular analysis (n= 122)** |  |
| *FLT3 ITD* | 17 (13.8) |
| *RUNX1-RUNX1T1* | 53 (43.1) |
| *CBFB-MYH11* | 6 (4.9) |
| *NPM1* | 5 (4.1) |
| Negative | 41 (33.6) |
| **ELN risk stratification\*\* (n=134)** |  |
| Good | 67 (50) |
| Intermediate | 37 (27.6) |
| Poor | 30 (22.4) |
| **Complete remission (n=143)** |  |
| Achieved | 104 (72.7%) |
| Not achieved | 39 (27.3%) |

\* t(8;21)= t(8;21)(q22;q22) *RUNX1-RUNX1T1*; inv(16)= inv(16)(p13.1;q22) *CBFB-MYH11*
\*\*ELN risk stratification was done using both cytogenetics and molecular markers in 134 patients. However, for 12 patients risk stratification was done with only cytogenetics and in 20 patients, it was done by only molecular analysis.
Median values were reported with range
n: number of patients; AML: Acute Myeloid Leukemia; Normal cytogenetics: 46,XX/46,XY; Failed cytogenetics: metaphases could not be isolated; *FLT3 ITD*: FMS-like tyrosine kinase internal tandem duplication; *RUNX1-RUNX1T1*: runt-related transcription factor 1-RUNX1 partner transcriptional co-repressor 1 fusion transcript; *CBFB-MYH11*: core binding factor beta-myosin heavy chain 11 fusion transcript; *NPM1*: Nucleophosmin 1; ELN: European LeukemiaNet.

### Identification of DEGs in paediatric AML based on mtDNA copy number

We identified 898,769, and 953 significantly dysregulated transcripts in AMLCN_H, AMLCN_I and AMLCN_L groups respectively by whole transcriptome sequencing as represented in volcano plots (Figure3). Majority of genes were found significantly downregulated in all three groups whereas the number of dysregulated genes were higher in AMLCN_H group compared to other two groups. A total of 351 DEGs (59 upregulated and 292 downregulated) were identified in AMLCN_H. Similarly, AMLCN_I and AMLCN_L groups had 290 (66 upregulated and 224 downregulated) and 332 (47 upregulated and 285 downregulated) DEGs respectively as compared to controls.

**Figure 3:**
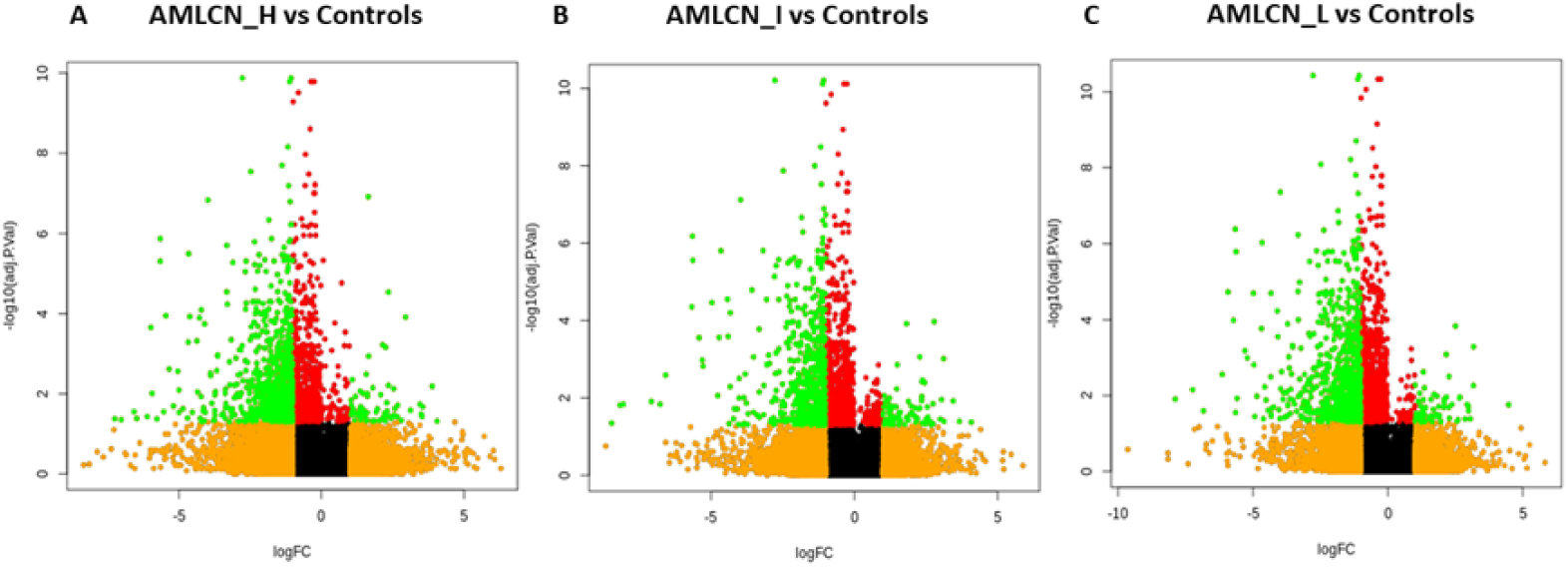
Volcano plots representing all dysregulated transcripts among AMLCN_H (A), AMLCN_I (B) and AMLCN_L (C) groups compared to controls. Green dots represent significantly differentially expressed transcripts in the three subgroups of AML compared to controls. High AMLCN_H (more than 75^th^ percentile); intermediate AMLCN_I (50^th^ to 75^th^ percentile) and low AMLCN_L (lower than 50^th^ percentile) relative mitochondrial DNA copy number

### Identification of mitochondria-related DEGs, hub genes and selection of genes for validation

Out of all DEGs, 78, 58, and 71 mitochondria-related DEGs were identified in AMLCN_H, AMLCN_I and AMLCN_L groups respectively. Among them, 35 genes were common in all three subgroups, whereas 18, 12 and 14 mitochondria-related DEGs were exclusively present in AMLCN_H, AMLCN_I and AMLCN_L groups respectively (Figure 4). In AMLCN_H, AMLCN_I and AMLCN_L groups, we identified 17, 18 and 17 hub genes respectively using CytoHubba analysis, of which eight were common among all subgroups (Table S3, Figure 5). Furthermore, using MCODE analysis, clusters with maximum scores were generated and seed gene was determined in the three groups (Figure 6). *MMP9* was identified as seed node with maximum MCODE score in both AMLCN_H and AMLCN_I group (Table S4). Based on the mitochondrial compartment score, CytoHubba and MCODE analyses, a total of 20 DEGs were selected for further validation (Table 2).

**Figure 4:**
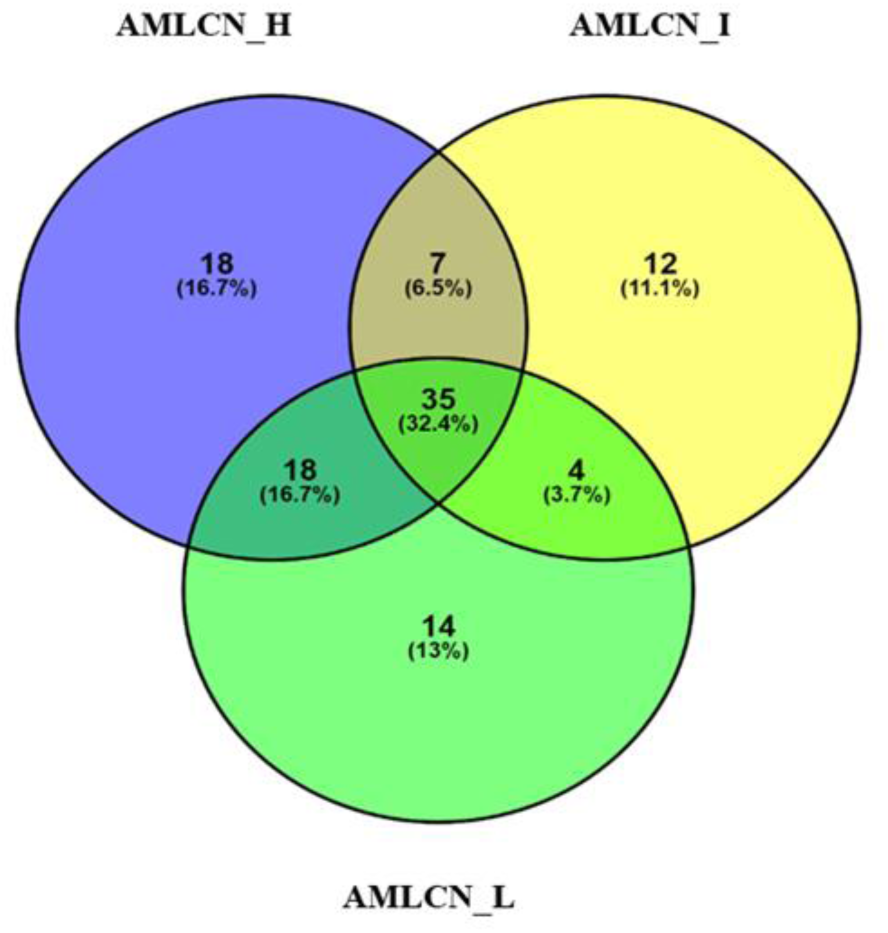
Venn diagram representing mitochondrial associated differentially expressed genes (DEGs) among three groups of AML (AMLCN_H; AMLCN_I and AMLCN_L). High AMLCN_H (more than 75^th^ percentile); intermediate AMLCN_I (50^th^ to 75^th^ percentile) and low AMLCN_L (lower than 50^th^ percentile) relative mitochondrial DNA copy number

**Figure 5:**
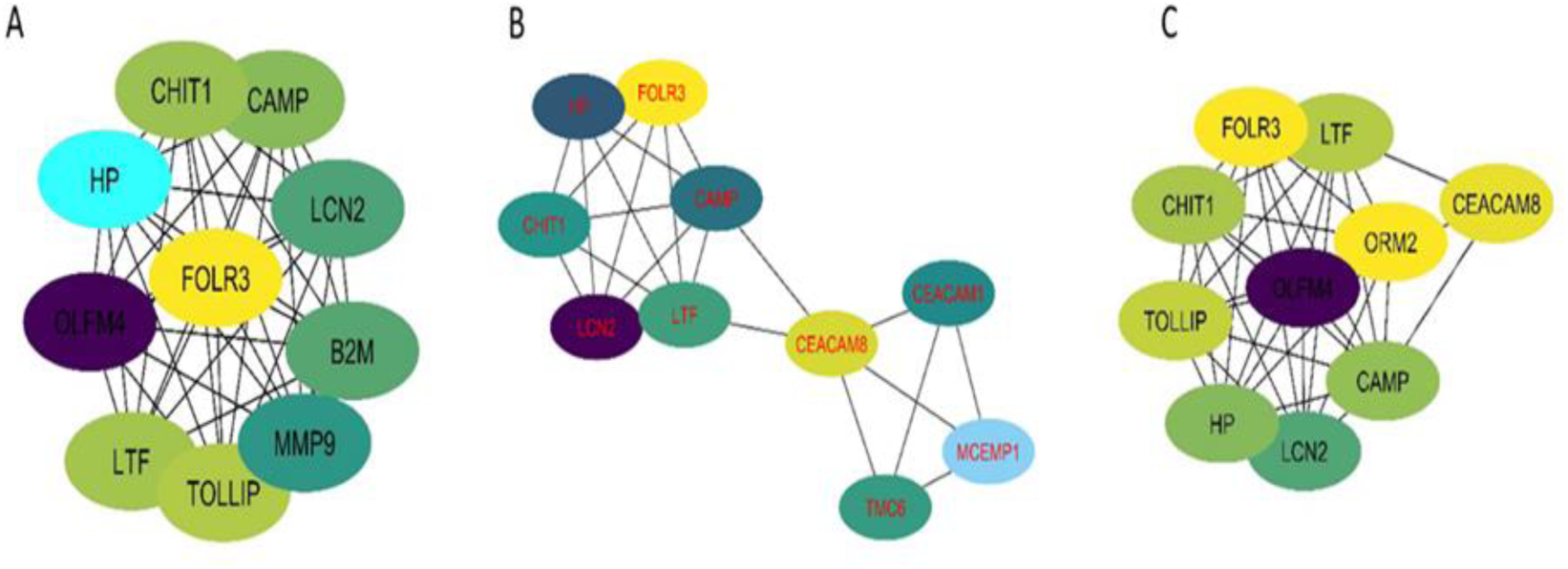
Hub gene based on Maximal Clique Centrality (MCC) (A): AMLCN_H; (B) AMLCN_I; (C) AMLCN_L. High AMLCN_H (more than 75^th^ percentile); intermediate AMLCN_I (50^th^ to 75^th^ percentile) and low AMLCN_L (lower than 50^th^ percentile) relative mitochondrial DNA copy number

**Figure 6:**
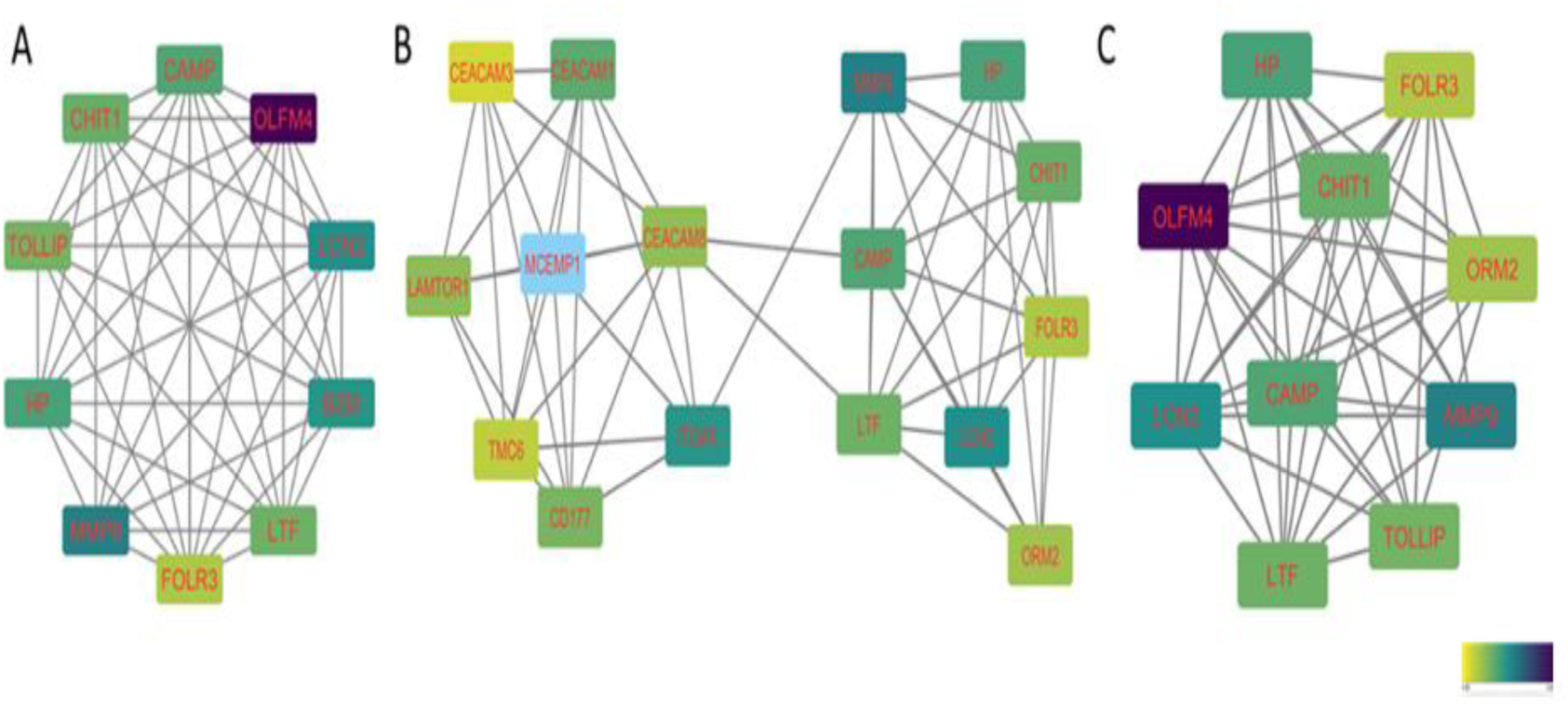
MCODE (molecular complex detection) analysis showing most interactive clusters among A: (A): AMLCN_H; (B) AMLCN_I; (C) AMLCN_L. High AMLCN_H (more than 75^th^ percentile); intermediate AMLCN_I (50^th^ to 75^th^ percentile) and low AMLCN_L (lower than 50^th^ percentile) relative mitochondrial DNA copy number

**Table 2:** List of genes selected for validation among all the groups based on mitochondrial compartment score, *Cytohubba* and *MCODE*

| S no. | Gene name | Compartment mitochondria score (0-5) | Groupwise expression | Expression in RNA seq data |
| --- | --- | --- | --- | --- |
| 1 | *SLC25A3* | 5 | AMLCN_H | Upregulation |
| 2 | *LONP1* | 5 | AMLCN_H | Downregulation |
| 3 | *SDHC* | 5 | AMLCN_H | Upregulation |
| 4 | *GNB2L1/RACK1* | 4.2 | AMLCN_H | Upregulation |
| 5 | *FASTKD1* | 5 | AMLCN_I | Upregulation |
| 6 | *MRPL51* | 5 | AMLCN_I | Downregulation |
| 7 | *ATP5J* | 5 | AMLCN_I | Upregulation |
| 8 | *FASLG* | 2.64 | AMLCN_L | Downregulation |
| 9 | *CLIC1* | 3.12 | AMLCN_L | Upregulation |
| 10 | *HRK* | 5 | Common in all AML samples | Downregulation |
| 11 | *ALAS2* | 5 | Common in all AML samples | Downregulation |
| 12 | *SLC25A21* | 5 | Common in all AML samples | Downregulation |
| 13 | *CYP11B1* | 4.35 | Common in all AML samples | Downregulation |
| 14 | *GLUD1* | 5 | Common in AMLCN_H & AMLCN_I | Upregulation |
| 15 | *SLC25A29* | 5 | Common in AMLCN_H & AMLCN_I | Upregulation |
| 16 | *LIG1* | 3.6 | Common in AMLCN_H & AMLCN_I | Downregulation |
| 17 | *SNCA* | 4.6 | Common in AMLCN_L & AMLCN_H | Downregulation |
| 18 | *DHFR* | 4.7 | Common in AMLCN_L & AMLCN_H | Downregulation |
| 19 | *MMP9* | 2.8 | Hub gene and Seed gene in MCODE cluster of AMLCN_H & AMLCN_I | Downregulation |
| 20 | *OLFM4* | 5 | MCODE cluster gene | Downregulation |

All the genes selected for validation are listed in the table. Selection of genes were based on their mitochondrial compartment score as determined by *Cytoscape*. *SLC25A3*, *LONP1*, *SDHC*, *GNB2L1/RACK1* were selected exclusively from AMLCN\_H subgroup. Three genes (*FASTKD1*, *MRPL51* AND *ATP6J*) belongs to AMLCN\_I subgroup and two genes (*FASLG* & *CLIC1*) were selected from AMLCN\_L subgroup. Furthermore, three genes (*SLC25A29*, *GLUD1* and *LIG1*) which were common among AMLCN\_H and AMLCN\_I and two genes, *DHFR* and *SNCA* were selected from the common genes among AMLCN\_H and AMLCN\_L cohort. *MMP9* and *OLFM4* were selected based on the Hub gene and MCODE analysis.

### Validation of selected DEGs, comparison with TCGA database

In the validation cohort of 143 AML patients, the expression of *SLC25A3*, *SDHC*, *RACK1/GNB2L1*, *FASTKD1*, *ATP5J*, *CLIC1*, *GLUD1,* and *SLC25A29* were found to be significantly upregulated (Figure 7, Table 3) while *FASLG*, *HRK*, *ALAS2*, *SLC25A21*, *CYP1B1*, *SNCA*, *MMP9,* and *OLFM4* were significantly downregulated (Figure 8, Table 3) compared to controls. Two selected genes, *LIG1* and *MRPL51* did not show significant dysregulation while *LONP1* had a reverse expression in the validation compared to transcriptomic expression profile. Upon comparison with TCGA dataset of adult AML patients, similar dysregulation was observed for *ALAS2, SLC25A21* and *SLC25A29* genes while a reverse expression pattern was observed for *ATP5J* and *CLIC1* genes; none of the other genes showed significant dysregulation in the TCGA dataset (Table 3).

**Figure 7:**
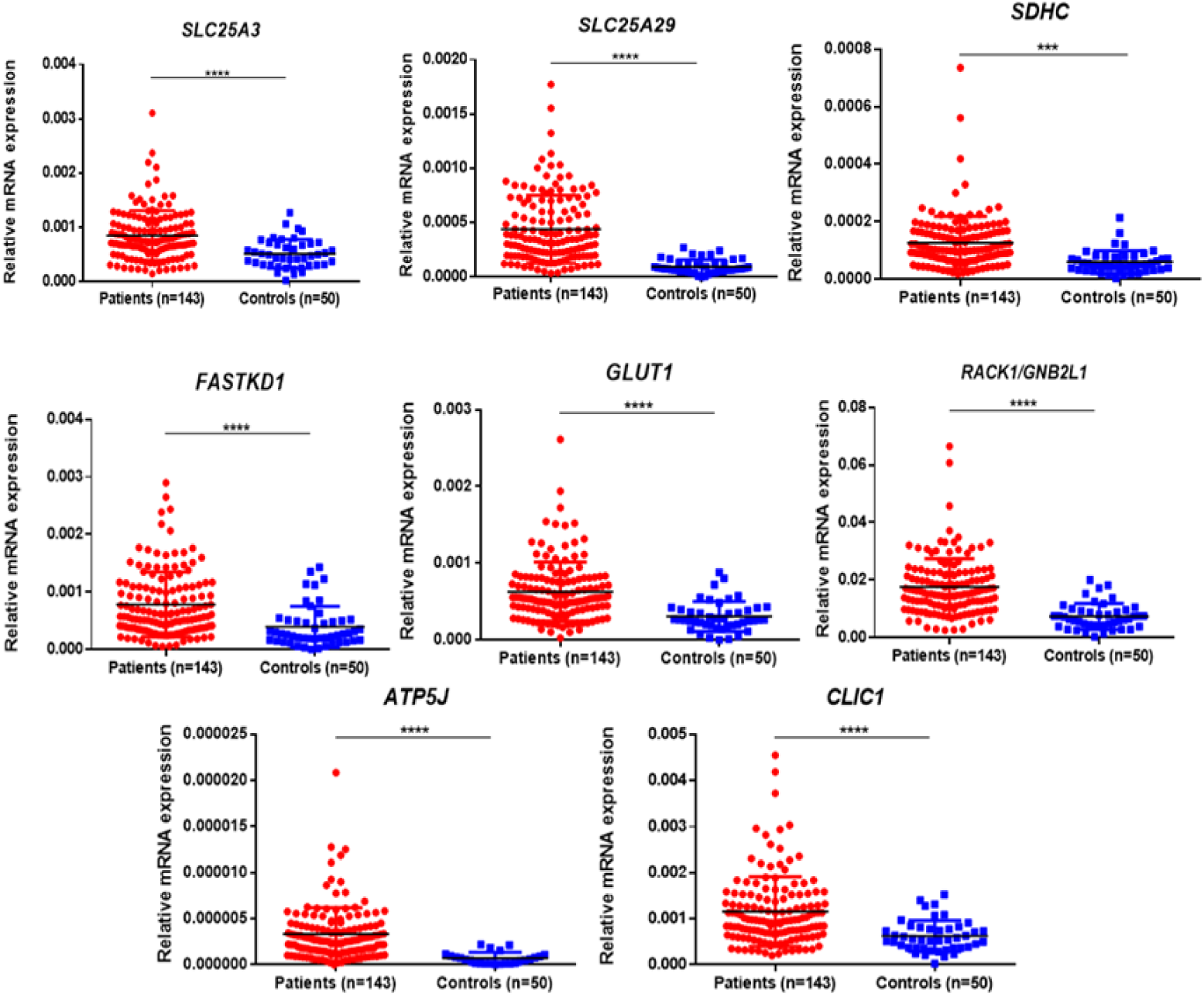
Validation of selected upregulated differentially expressed genes (DEGs) in patients as compared to controls. *SLC25A3, SLC25A29, SDHC, FASTKD1, GLUD1, RACK1, ATP5J* and *CLIC1* were significantly upregulated in pediatric AML patients (n=143) compared to controls (n=50). *: P<0.05; **: P< 0.01; ***: P<0.001; ****: P<0.0001.

**Figure 8:**
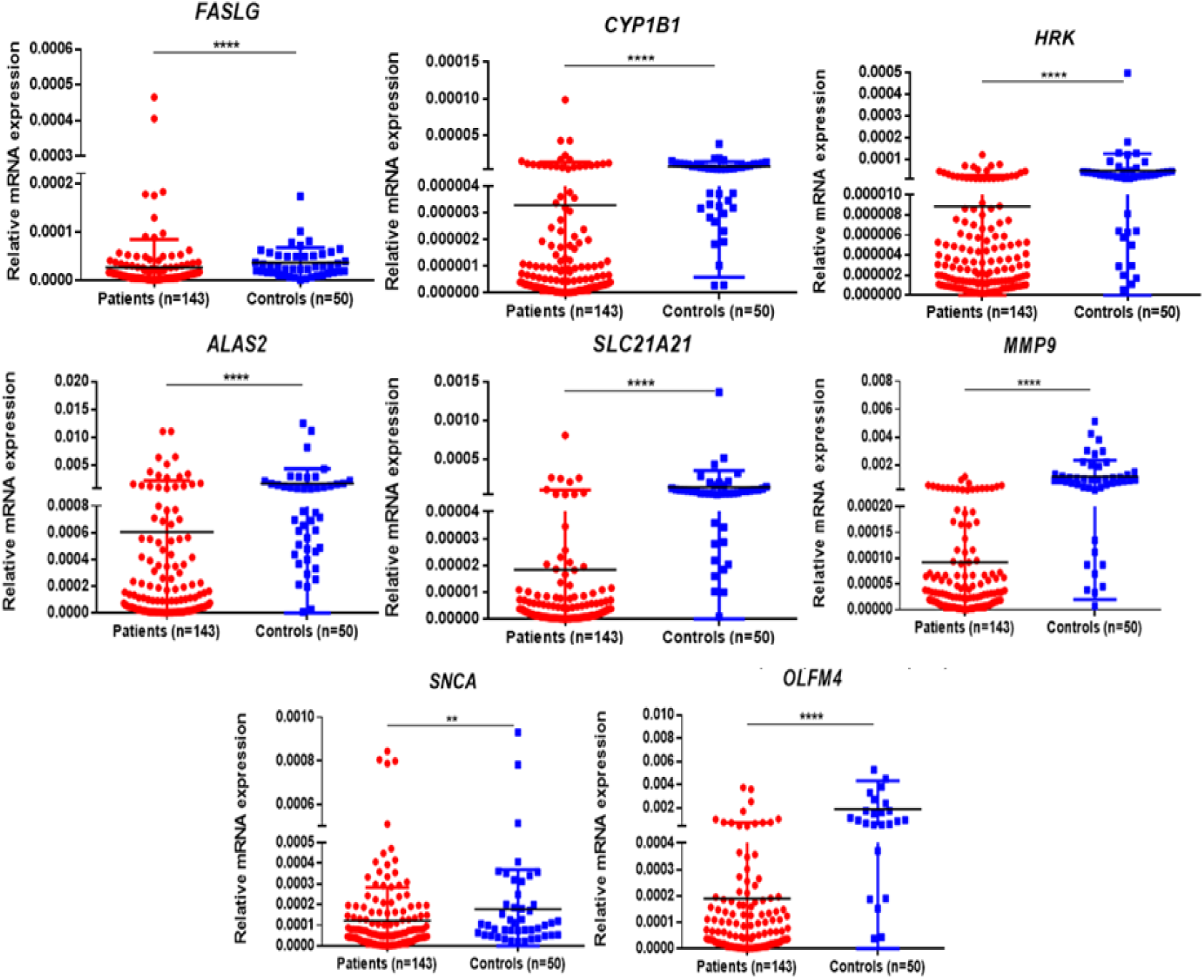
Validation of selected downregulated differentially expressed genes (DEGs) in patients as compared to controls. *FASLG, CYP1B1, HRK, ALAS2, SLC25A21, MMP9, SNCA* and *OLFM4* were significantly downregulated in pediatric AML patients (n=143) compared to controls (n=50) *: P<0.05; **: P< 0.01; ***: P<0.001; ****: P<0.0001

**Table 3:**
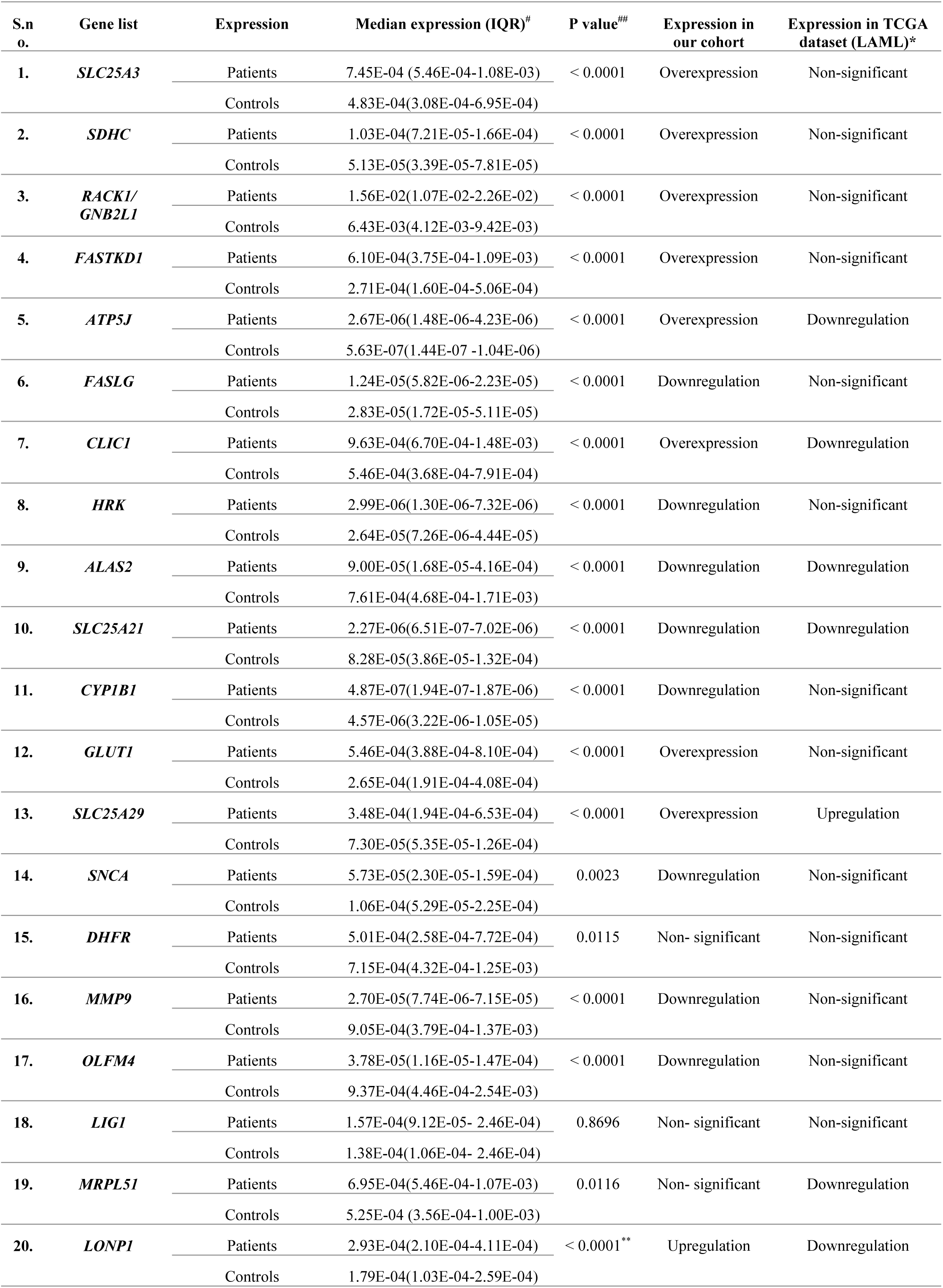
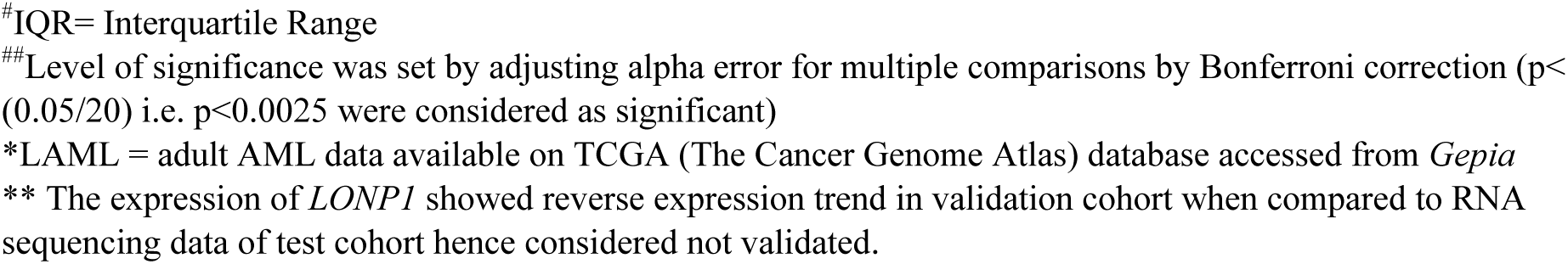
Median expression of validated genes in patients (n=143) compared to controls(n=50) and their comparison with TCGA LAML dataset(n=179)

| S.n o. | Gene list | Expression | Median expression (IQR)<sup>#</sup> | P value<sup>##</sup> | Expression in our cohort | Expression in TCGA dataset (LAML)* |
| --- | --- | --- | --- | --- | --- | --- |
| 1. | *SLC25A3* | Patients | 7.45E-04 (5.46E-04-1.08E-03) | < 0.0001 | Overexpression | Non-significant |
|  |  | Controls | 4.83E-04(3.08E-04-6.95E-04) |  |  |  |
| 2. | *SDHC* | Patients | 1.03E-04(7.21E-05-1.66E-04) | < 0.0001 | Overexpression | Non-significant |
|  |  | Controls | 5.13E-05(3.39E-05-7.81E-05) |  |  |  |
| 3. | *RACK1/*<br>*GNB2L1* | Patients | 1.56E-02(1.07E-02-2.26E-02) | < 0.0001 | Overexpression | Non-significant |
|  |  | Controls | 6.43E-03(4.12E-03-9.42E-03) |  |  |  |
| 4. | *FASTKD1* | Patients | 6.10E-04(3.75E-04-1.09E-03) | < 0.0001 | Overexpression | Non-significant |
|  |  | Controls | 2.71E-04(1.60E-04-5.06E-04) |  |  |  |
| 5. | *ATP5J* | Patients | 2.67E-06(1.48E-06-4.23E-06) | < 0.0001 | Overexpression | Downregulation |
|  |  | Controls | 5.63E-07(1.44E-07 -1.04E-06) |  |  |  |
| 6. | *FASLG* | Patients | 1.24E-05(5.82E-06-2.23E-05) | < 0.0001 | Downregulation | Non-significant |
|  |  | Controls | 2.83E-05(1.72E-05-5.11E-05) |  |  |  |
| 7. | *CLIC1* | Patients | 9.63E-04(6.70E-04-1.48E-03) | < 0.0001 | Overexpression | Downregulation |
|  |  | Controls | 5.46E-04(3.68E-04-7.91E-04) |  |  |  |
| 8. | *HRK* | Patients | 2.99E-06(1.30E-06-7.32E-06) | < 0.0001 | Downregulation | Non-significant |
|  |  | Controls | 2.64E-05(7.26E-06-4.44E-05) |  |  |  |
| 9. | *ALAS2* | Patients | 9.00E-05(1.68E-05-4.16E-04) | < 0.0001 | Downregulation | Downregulation |
|  |  | Controls | 7.61E-04(4.68E-04-1.71E-03) |  |  |  |
| 10. | *SLC25A21* | Patients | 2.27E-06(6.51E-07-7.02E-06) | < 0.0001 | Downregulation | Downregulation |
|  |  | Controls | 8.28E-05(3.86E-05-1.32E-04) |  |  |  |
| 11. | *CYP11B1* | Patients | 4.87E-07(1.94E-07-1.87E-06) | < 0.0001 | Downregulation | Non-significant |
|  |  | Controls | 4.57E-06(3.22E-06-1.05E-05) |  |  |  |
| 12. | *GLUT1* | Patients | 5.46E-04(3.88E-04-8.10E-04) | < 0.0001 | Overexpression | Non-significant |
|  |  | Controls | 2.65E-04(1.91E-04-4.08E-04) |  |  |  |
| 13. | *SLC25A29* | Patients | 3.48E-04(1.94E-04-6.53E-04) | < 0.0001 | Overexpression | Upregulation |
|  |  | Controls | 7.30E-05(5.35E-05-1.26E-04) |  |  |  |
| 14. | *SNCA* | Patients | 5.73E-05(2.30E-05-1.59E-04) | 0.0023 | Downregulation | Non-significant |
|  |  | Controls | 1.06E-04(5.29E-05-2.25E-04) |  |  |  |
| 15. | *DHFR* | Patients | 5.01E-04(2.58E-04-7.72E-04) | 0.0115 | Non- significant | Non-significant |
|  |  | Controls | 7.15E-04(4.32E-04-1.25E-03) |  |  |  |
| 16. | *MMP9* | Patients | 2.70E-05(7.74E-06-7.15E-05) | < 0.0001 | Downregulation | Non-significant |
|  |  | Controls | 9.05E-04(3.79E-04-1.37E-03) |  |  |  |
| 17. | *OLFM4* | Patients | 3.78E-05(1.16E-05-1.47E-04) | < 0.0001 | Downregulation | Non-significant |
|  |  | Controls | 9.37E-04(4.46E-04-2.54E-03) |  |  |  |
| 18. | *LIG1* | Patients | 1.57E-04(9.12E-05- 2.46E-04) | 0.8696 | Non- significant | Non-significant |
|  |  | Controls | 1.38E-04(1.06E-04- 2.46E-04) |  |  |  |
| 19. | *MRPL51* | Patients | 6.95E-04(5.46E-04-1.07E-03) | 0.0116 | Non- significant | Downregulation |
|  |  | Controls | 5.25E-04 (3.56E-04-1.00E-03) |  |  |  |
| 20. | *LONPI* | Patients | 2.93E-04(2.10E-04-4.11E-04) | < 0.0001** | Upregulation | Downregulation |
|  |  | Controls | 1.79E-04(1.03E-04-2.59E-04) |  |  |  |

#IQR= Interquartile Range ##Level of significance was set by adjusting alpha error for multiple comparisons by Bonferroni correction ( $p < 0.05/20$ ) i.e. $p < 0.0025$ were considered as significant)
\*LAML = adult AML data available on TCGA (The Cancer Genome Atlas) database accessed from *Gepia* \*\* The expression of *LONPI* showed reverse expression trend in validation cohort when compared to RNA sequencing data of test cohort hence considered not validated.

### Mitochondria-related DEGs and mtDNA copy number

On univariable analysis, increased mtDNA copy number was significantly associated with poor event free survival (HR= 2.14; 95%CI (1.39-3.29); p=0.001) and overall survival (HR= 2.77; 95% CI (1.70-4.59); p<0.001) (Figure 9). In patients with increased mtDNA copy number, expression of *SLC25A3*, *SDHC*, *RACK1/GNB2L1* and *FASTKD1,* were significantly higher compared to those with low mtDNA copy number (Figure 10).Exclusive elevated expression of these 4 genes were also observed in transcriptome of samples with high/intermediate mtDNA copy number(AMLCN_H/AMLCN_I) compared to low mtDNA copy number (AMLCN_L).On correlation analysis, these 4 genes along with 2 other genes *CLIC1* and *ATP5J* showed significant positive correlation with mtDNA copy number(Table 4).

**Figure 9:**
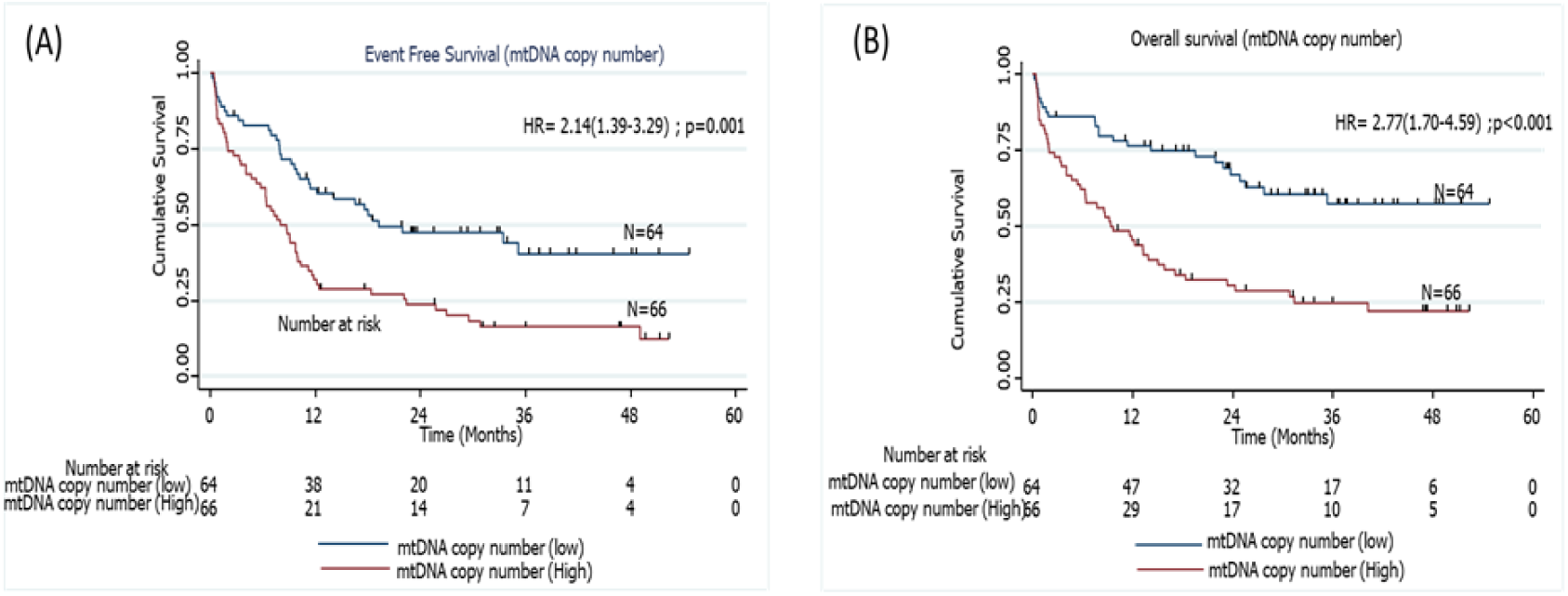
Kaplan Meier curves representing association of mtDNA copy number with (A) event free survival and (B) overall survival of pediatric AML patients

**Figure 10:**
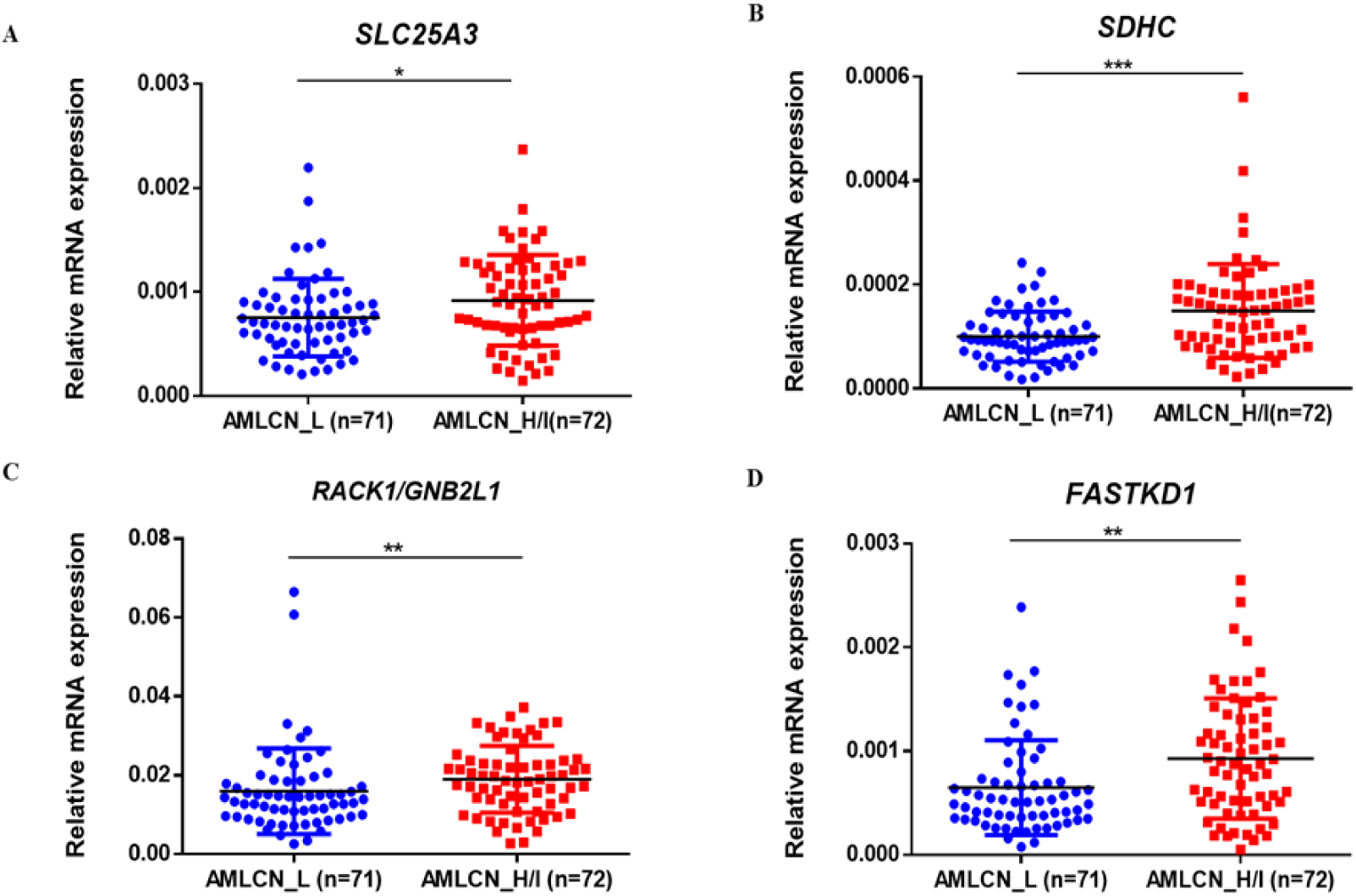
Subgroup analysis of acute myeloid leukemia showing exclusive deregulation of validated genes among AMLCN_L and AMLCN_H group. The expression of *SLC25A3*(A)*, SDHC*(B)*, RACK1*(C) and *FASTKD1*(D) were significantly higher in AMLCN_H group as compared to AMLCN_L

**Table 4:**
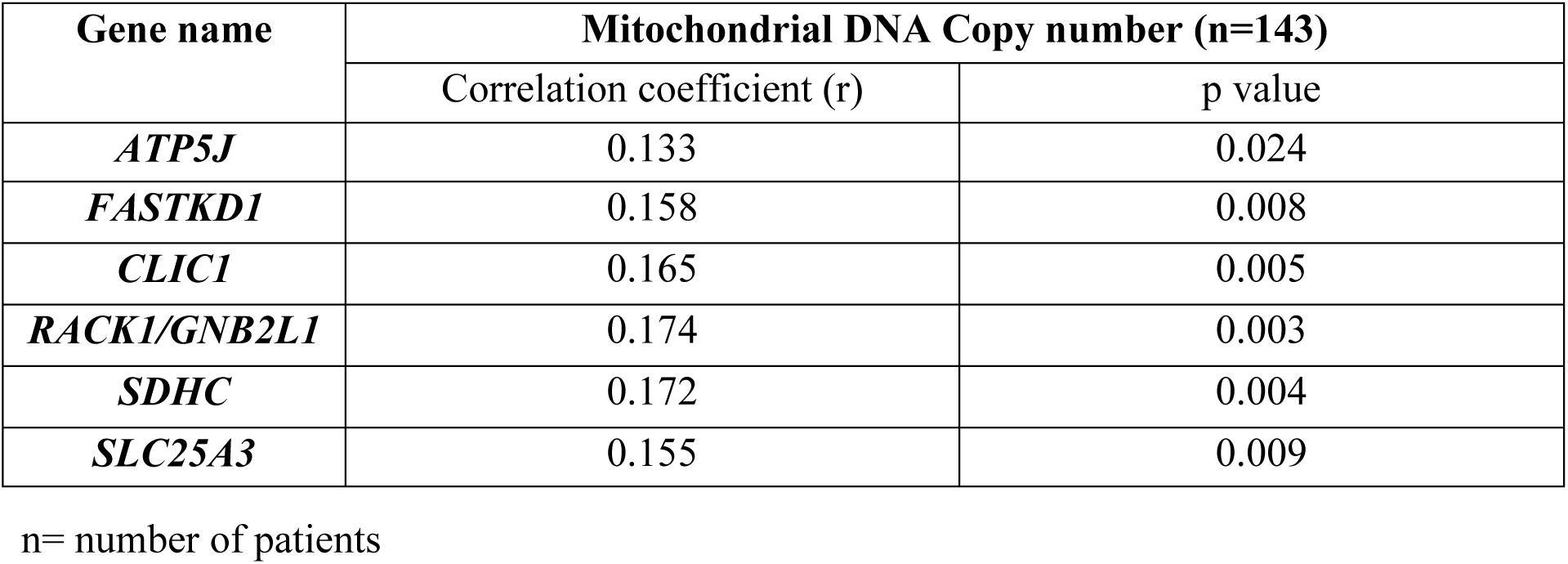
List of genes showing significant correlation with mitochondrial DNA copy number using Pearson’s correlation

| Gene name | Mitochondrial DNA Copy number (n=143) |  |
| --- | --- | --- |
|  | Correlation coefficient (r) | p value |
| *ATP5J* | 0.133 | 0.024 |
| *FASTKD1* | 0.158 | 0.008 |
| *CLIC1* | 0.165 | 0.005 |
| *RACK1/GNB2L1* | 0.174 | 0.003 |
| *SDHC* | 0.172 | 0.004 |
| *SLC25A3* | 0.155 | 0.009 |

n= number of patients

### Predictive ability of expression of validated DEGs on survival outcome and establishment of the prognostic gene signature

On multivariable analysis, upregulated expression of 2 genes, *SDHC* (HR 1.29; 95% CI (1.14-1.41); p<0.001) and*CLIC1*(HR 1.20; 95% CI (1.04-1.38); p=0.013), and downregulation of *SLC25A29*(HR 0.88; 95% CI (0.83-0.93); p<0.001) were found to be independently predictive of worse OS (Table 5) and they were included for the development of a prognostic gene signature model. All these 3 genes (*SDHC, CLIC1, SLC25A29*) satisfied internal validation by bootstrapping (Table S5), and were finally selected for prognostic model building. Beta coefficient of each of the variables were used for calculation of risk score as follows:

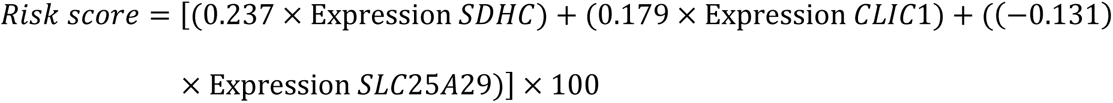

**Table 5:** Impact of expression of individual genes and overall risk score on overall survival and event free survival of the test cohort (pediatric cohort) and overall survival in validation cohort (TCGA adult LAML cohort)

| Gene Name | Overall survival (Pediatric cohort)<br>n=143 |  | Overall survival (TCGA adult LAML cohort)<br>n=179 |  | Event Free Survival (Pediatric cohort)<br>n=143 |  |
| --- | --- | --- | --- | --- | --- | --- |
|  | Hazard Ratio (95% CI) | P value | Hazard Ratio (95% CI) | P value | Hazard Ratio (95% CI) | P value |
| *SDHC* | 1.29(1.14-1.41) | <0.001 | 0.994(0.941-1.050) | 0.826 | 1.225(1.100-1.363) | <0.001 |
| *SLC25A29* | 0.88(0.83-0.93) | <0.001 | 0.988(0.981-0.996) | 0.003 | 0.905(0.860-0.952) | <0.001 |
| *CLIC1* | 1.20(1.04-1.38) | 0.013 | 1.002(1.00-1.004) | 0.069 | 1.136(0.984-1.312) | 0.082 |
| **Risk Score** | 1.010(1.007-1.014) | <0.001 | 1.011(1.002-1.021) | 0.019 | 1.008(1.001-1.012) | <0.001 |

CI: Confidence interval; n= number of patients; TCGA The cancer genome atlas; LAML Adult AML dataset

The formula was used to calculate risk score of all the patients. Risk score median value (10.382) was taken as the cut-off for subgrouping patients into high-risk and low risk group. Patients with high-risk scores (≥10.382) had inferior OS (HR 1.010; 95% CI (1.007-1.014): p<0.001) compared to those with low-risk score (<10.382) (Figure 11A). Harrel’s C-index of the prognostic model was 0.675. The timed AUC of the risk score for 12 months and 18 months survival was 0.747 and 0.736 respectively (Figure 11(B, C).

**Figure 11:**
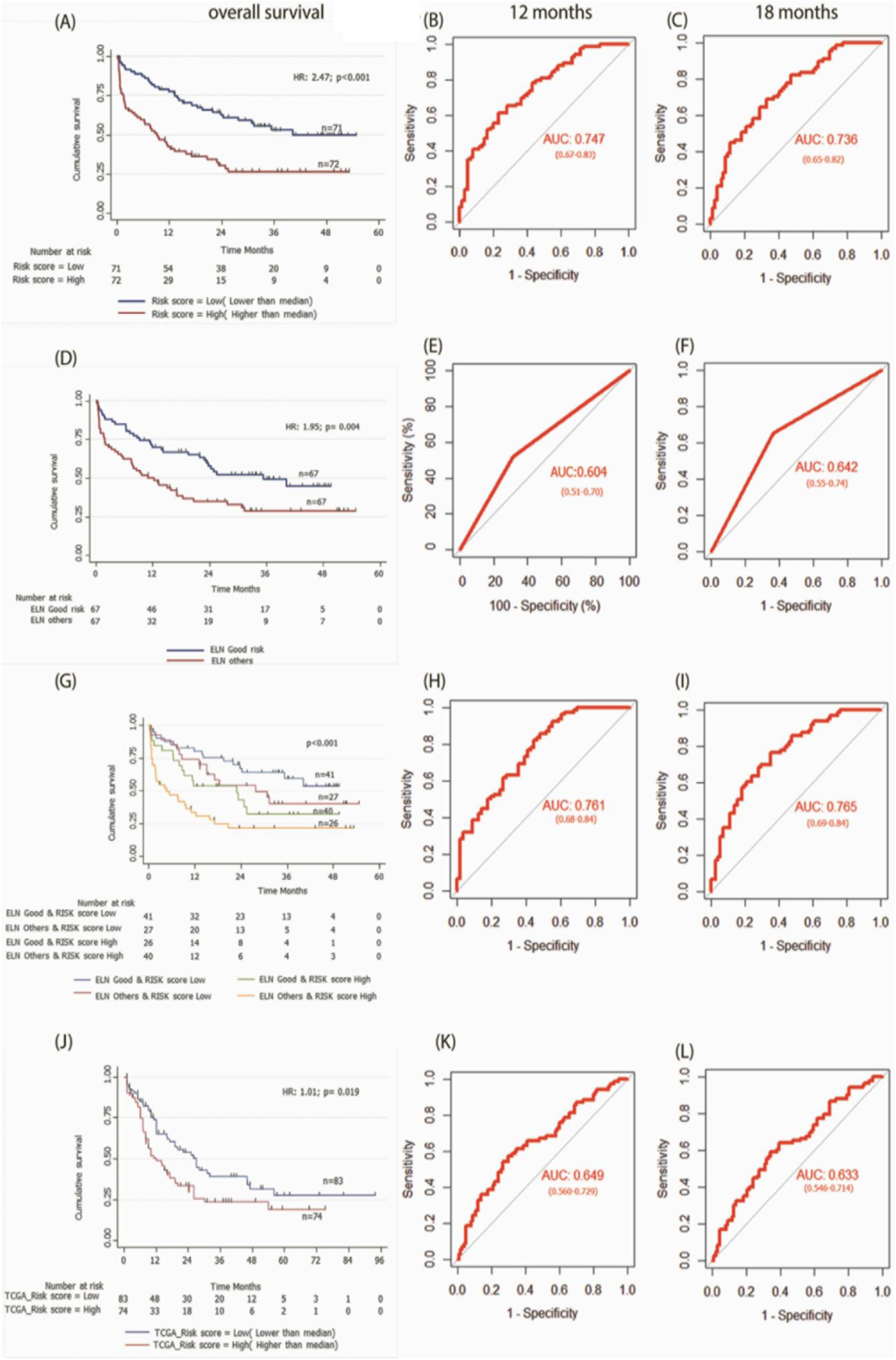
Kaplan Meier curves representing association of mtDNA copy number with (A) event free survival and (B) overall survival of pediatric AML patients **Figure 11.A 3-gene based gene signature stratifies survival in pediatric and adult AML patients along with clinically established European LeukemiaNet (ELN) risk categories.** (A) Kaplan Meier estimates of overall survival in pediatric AML patient’s subgroup into high Risk-score and low Risk-score. (B) and (C) AUC curves quantify the ability of our 3-gene based risk score to predict outcome in individual patients (specificity and sensitivity) within the first 12 months(B) and 18 months(C) of treatment initiation respectively. (D) Kaplan Meier estimates of overall survival in pediatric AML patient’s subgroup into ELN good risk and ELN intermediate or poor risk categories. (E) and (F) AUC curves quantify the ability of ELN risk categories to predict outcome in individual patients (specificity and sensitivity) within the first 12 months (E) and 18 months(F) of treatment initiation respectively. (G) Kaplan Meier estimates of overall survival in pediatric AML patient’s subgroup by combining ELN risk categories with our 3 gene-based risk score. (H) and (I) AUC curves quantify the ability of combined model of ELN risk categories and our 3 gene-based risk score to predict outcome in individual patients (specificity and sensitivity) within the first 12 months(H) and 18 months(I) of treatment initiation respectively. (J) Kaplan Meier estimates of overall survival in external adult The Cancer Genome Atlas (TCGA) AML patient’s subgroup into high Risk-score and low Risk-score using our 3 gene-based gene signature model. (K) and (L) AUC curves quantify the ability of our 3-gene based risk score to predict outcome in individual patients of TCGA adult AML datasets (specificity and sensitivity) within the first 12 months(K) and 18 months(L) of treatment initiation respectively. AUC = 1.0 would denote perfect prediction, and AUC = 0.5 would denote no predictive ability.

### Association of gene signature-based risk score with event free survival (EFS)

On multivariable Cox regression analysis, upregulation of *SDHC* (HR 1.225; 95% CI (1.100-1.363); p<0.001) and downregulation of *SLC25A29* (HR 0.905; 95% CI (0.860-0.952); p<0.001) were also predictive of worse EFS. We also found that patients with high-risk score had significantly lower EFS as compared to low-risk score patients (HR 1.008; 95% CI (1.001-1.012); p<0.001) (Table 5). Harrel’s C-index of prognostic model was 0.626. The timed AUC of the risk score for 12 months and 18 months EFS was 0.617 and 0.612 respectively

### Impact of baseline clinical features on survival outcome and association with gene signature model

On univariable Cox regression analysis, ELN intermediate/poor risk and absence of chloroma were significantly associated with inferior OS and only ELN category came out to be an independent prognostic factor in multivariable analysis (Table 6, Figure 11D). Furthermore, on multivariable analysis, both the ELN risk category (p=0.040) and risk score (p<0.001) were found to be independent prognostic factors for OS. We also performed multivariable analysis including mtDNA copy number and observed that all three factors i.e. risk score (p<0.001), ENA risk categories(p=0.012) and mtDNA copy number(p=0.012) were independent prognostic factors for OS (Table S6).

**Table 6:** Univariable and multivariable analysis of clinical features with overall survival

| Variables (n=143) | Categories (n) | Univariable analysis |  |  | Multivariable analysis |  |
| --- | --- | --- | --- | --- | --- | --- |
|  |  | Median | Hazard (95% CI) | p value | Hazard (95% CI) | p value |
| **Age(years)** | ≤10 (64) | 20.63 | 0.98(0.63-1.52) | 0.93 | - | - |
|  | >10(79) | 21.93 |  |  |  |  |
| **Gender** | Male (87) | 15.07 | 0.72(0.45-1.143) | 0.16 | - | - |
|  | Female (56) | 40.23 |  |  |  |  |
| **Total leukocyte count (/mm<sup>3</sup>)** | <50000(98) | 23.77 | 0.8(0.517-1.238) | 0.32 | - | - |
|  | >50000(45) | 13.88 |  |  |  |  |
| **Platelets (/μL)** | ≤ 50000(92) | 19.53 | 0.94(0.59-1.48) | 0.789 | - | - |
|  | >50000(51) | 23.33 |  |  |  |  |
| **Haemoglobin(g/dl)** | ≤8(95) | 13.87 | 0.73(0.47-1.12) | 0.16 | - | - |
|  | >8(48) | 24.87 |  |  |  |  |
| **Fever** | Negative (26) | Not reached | 1.52(0.82-2.83) | 0.18 | - | - |
|  | Positive (112) | 19.53 |  |  |  |  |
| **Chloroma (n=132)** | Negative (105) | 20.63 | 0.56(0.29-1.07) | 0.083 | 1.45(0.74-2.84) | 0.27 |
|  | Positive (27) | Not reached |  |  |  |  |
| **ELN Risk group (n=134)** | Good (67) | 40.23 | 1.95(1.23-3.09) | 0.004 | 0.59(0.36-0.98) | 0.041 |
|  | Others (67) | 12.27 |  |  |  |  |

CI: Confidence interval; ELN: European LeukemiaNet.

### Impact of combined clinical and gene signature model on survival outcome of the cohort

To compare the predictive ability of our gene signature risk score and ELN risk stratification on OS of AML patients, a time dependent AUC was constructed. Harrel’s C-index of the model was 0.59 and the timed AUC of ELN risk category on 12 months and 18 months survival was 0.60 and0.64 respectively (Figure 11E, F). We combined the ELN risk strategy with our risk score and calculated the predictive ability of the model. The Harrel’s C-index of the model was 0.688 and the timed AUC of combining ELN risk strategy with gene signature risk score for 12 months and 18 months was 0.761 and 0.765 respectively (Figure 11H and 2I).

### Association of gene signature risk score on disease characteristics

We found that a high-risk score was significantly associated with poor risk cytogenetics(p=0.021), absence of RUNX1-RUNX1T1 translocation (p=0.027) and ELN intermediate/poor risk group (p=0.016). Furthermore, the proportion of patients achieving CR was significantly higher in the low-risk group as compared to the high-risk group(p=0.017) (Table 7). On subgroup analysis, it was observed that the mitochondria-related gene signature risk score category was significantly predictive of survival outcome across all clinically relevant subgroups except in those with intermediate/poor-risk karyotype (Figure 12).

**Figure 12:**
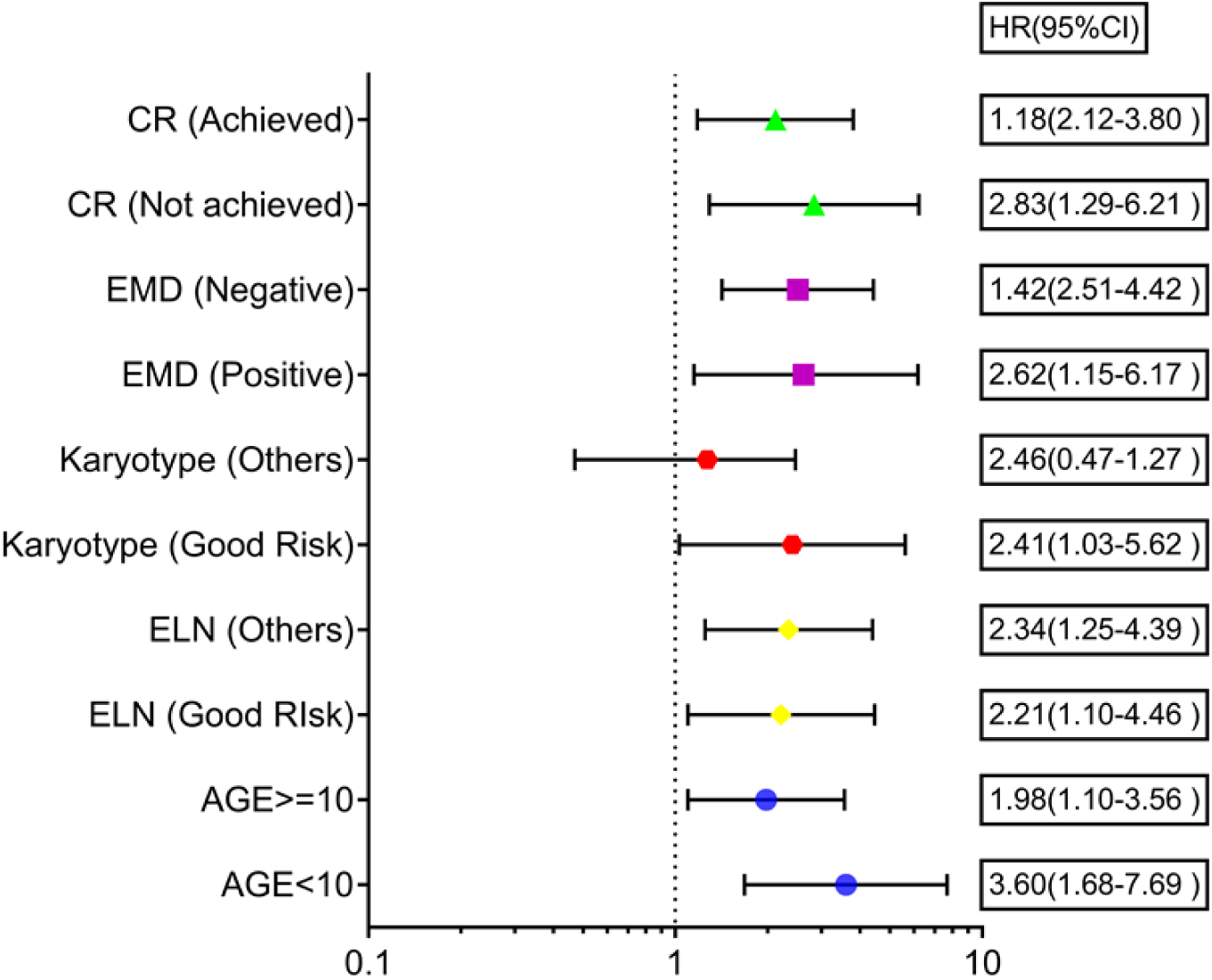
Forest plot showing the impact of mitochondrial prognostic gene signature risk score category on survival outcome in various clinically relevant subgroups of pediatric Acute myeloid leukemia

**Table 7:** Association of 3-gene risk score with clinical and demographic parameters

| Characteristics (n=143) | Risk score<br>Low (%)<br>(n=71) | Risk score<br>High (%)<br>(n=72) | $\chi^2$ | P value |
| --- | --- | --- | --- | --- |
| Age (years) |  |  |  |  |
| <10 Years (64) | 26(36.6) | 38(52.8) | 3.775 | 0.052 |
| ≥10 years (79) | 45(63.4) | 34(47.2) |  |  |
| Sex |  |  |  |  |
| Male (87) | 42(59.2) | 45(62.5) | 0.168 | 0.682 |
| Female (56) | 29(40.8) | 27(37.5) |  |  |
| Hyperleukocytosis, (>50×10<sup>3</sup>/μL) |  |  |  |  |
| TLC <50×10<sup>3</sup>/μL (98) | 53(74.6) | 45(62.5) | 2.446 | 0.118 |
| TLC≥50×10<sup>3</sup>/μL (45) | 18(25.4) | 27(37.5) |  |  |
| Fever(n=138) |  |  |  |  |
| Negative (26) | 17(24.6) | 9(13.0) | 3.033 | 0.082 |
| Positive (112) | 52(75.4) | 60(87.0) |  |  |
| Chloroma |  |  |  |  |
| Negative (116) | 57(80.3) | 59(81.9) | 1.781 | 0.182 |
| Positive (27) | 14(19.7) | 13(18.1) |  |  |
| Cytogenetics (n=130) * |  |  |  |  |
| Good Risk (50) | 32(54.2) | 18(32.7) | 5.349 | 0.021 |
| Others (64) | 27(45.8) | 37(67.3) |  |  |
| Molecular analysis (n= 122) ** |  |  |  |  |
| FLT3ITD |  |  |  |  |
| Negative (105) | 54(87.1) | 51(85.0) | 0.112 | 0.738 |
| Positive (17) | 8(12.9) | 9(15.0) |  |  |
| RUNX1-RUNX1T1 |  |  |  |  |
| Negative (69) | 29(46.8) | 40(66.7) | 4.911 | 0.027 |
| Positive (53) | 33(53.2) | 20(33.3) |  |  |
| CBFB-MYH11 # |  |  |  |  |
| Negative (113) | 58(93.5) | 55(91.7) | - | 1.00<sup>##</sup> |
| Positive (6) | 3(4.83) | 3(5.0) |  |  |
| NPM1 |  |  |  |  |
| Negative (117) | 61(98.4) | 56(93.3) | - | 0.203<sup>##</sup> |
| Positive (5) | 1(1.61) | 4(6.7) |  |  |

| ELN Risk stratification (n=134) *** |  |  |  |  |
| --- | --- | --- | --- | --- |
| Good Risk (67) | 41(60.3) | 26(39.4) | 5.852 | 0.016 |
| Intermediate and poor risk (67) | 27(39.7) | 40(60.6) |  |  |
| Complete remission (n=143) |  |  |  |  |
| Achieved (104) | 58(81.7) | 46(63.9) | 5.711 | 0.017 |
| Not achieved (39) | 13(18.3) | 26(36.1) |  |  |

\*Cytogenetics failed (n=16) and not done in n=13 cases.
\*\* Molecular analysis was not done in 19 cases; molecular mutation was absent in n=43 cases
#*CBFB-MYH11* mutation was assessed in n=119 cases

### Predictive ability of combined gene signature and ELN category model on survival outcome

The predictive ability of gene signature score along with ELN risk stratification on survival outcome of paediatric AML patients was also assessed. Patients with low gene signature score (low risk) belonging to ELN good risk category had significantly better survival outcome (Median OS: Not reached) and predicted 12-months (80%±6%), as well as 18-months (75%±7%) survival. Similarly, patients with high gene signature score (high risk) belonging to ELN intermediate/poor risk category had significantly inferior outcome (4.67 months (0-3.71)) with 12-months and 18-months predicted survival of 33% ± 8% and 25% ± 7% respectively. On the other hand, patients belonging to other groups (ELN intermediate/poor risk and low-risk; ELN good risk and high-risk score) had intermediate survival outcome (median survival of 27.77-22.90 months respectively) between the two other groups (Figure 11G; Table 8).

**Table 8:** Predictive ability of combined gene risk score group and ELN risk category on survival outcome in pediatric AML cohort

| Variables(n) | HR (95%CI) | P Value | P value (overall) | Median OS (months) | Predicted 12 months survival | Predicted 18 months survival |
| --- | --- | --- | --- | --- | --- | --- |
| **ELN Good risk & gene signature risk group Low (41)** | 1 | -- | <0.001 | Not reached | 80%±6% | 75%±7% |
| **ELN Others & gene signature risk group Low (27)** | 1.58 (0.78-3.21) | 0.20 | - | 27.77(±7.82) | 74% ±8% | 58% ±10% |
| **ELN Good risk & gene signature risk group High (26)** | 2.12 (1.06-4.26) | 0.034 | - | 22.90(±8.31) | 54% ±10% | 44% ±10% |
| **ELN Others & gene signature risk group High (40)** | 3.83 (2.07-7.07) | <0.001 | - | 4.67(±3.71) | 33% ± 8% | 25% ± 7% |

HR: Hazard Ratio; CI: Confidence interval; OS: overall survival; ELN: European LeukemiaNet; AML: Acute myeloid leukemia

### External validation of gene signature risk score in TCGA database

Using our risk calculation model, we calculated the risk score in TCGA dataset (n=179) and similarly, patients were further sub-grouped as high-risk score (higher than median) and low risk score (lower than median) based on the median value (43.434). Kaplan Meier analysis showed that patients with a high-risk score (≥43.434) had inferior OS (HR 1.01;95% CI (1.00-1.02); p<0.019) compared to those with a low-risk score (<43.434) (Figure 11J; Table 5). Along with this, poor risk karyotype patients had worse overall survival (HR 1.89; 95% CI (2.95-1.20); p=0.004) compared to patients with good risk or intermediate risk karyotype (7.03 vs 18.96months). On multivariable analysis karyotype (poor vs good risk/intermediate risk) and risk score were found to be independently predictive of (p=0.002; p=0.025 respectively) for worse OS. The timed AUC of risk score for 12-months and 18-months survival in the TCGA dataset were 0.64 and 0.63 respectively (Figure 11K and 11L) and Harrel’s C-index of the prognostic model was 0.600.

## Discussion

Our study is the first one to identify and validate mitochondria-related DEGs in paediatric AML along with determining their prognostic significance. In paediatric AML patients, we identified and validated 16 mitochondrial DEGs including 8 upregulated and 8 downregulated genes compared to controls. The dysregulated expression of these genes has been previously reported in the pathogenesis of various malignancies(20–23). However, they have not been studied in paediatric AML. Comparison with LAML dataset of TCGA cohort suggests that the mitochondria-related gene expression profile in paediatric AML is likely distinct. Elevated expression of genes like *SLC25A3, FASTKD1, SDHC, ATP5J*, which were observed for the first time in our cohort, are involved in mitochondrial energy metabolism(24–26). Genes like *FASLG, HRK* and *SNCA,* which were observed to be downregulated, also play role in prevention of mitochondrial damage and apoptosis inhibition in melanoma/medulloblastoma cell lines(27–29). Preliminary data suggests that downregulation of genes like *MMP9* and *OLFM4*, as observed in our cohort, may aid in AML progression(30, 31). The expression of *CYP1B1* is reported to be elevated in various malignancies, however, its expression is downregulated in early age leukaemia, as seen in our cohort(32). These findings suggest that the observed mitochondria-related DEGs likely play crucial role in disease progression in paediatric AML, which needs to be studied further mechanistically.

Enhanced mtDNA copy number has been previously reported to be play role on AML initiation, progression as well as predictive of survival outcomes(12, 33). Similar to our previous finding, we found that mtDNA copy number were significantly higher and independently predictive of worse survival outcome in this cohort of pediatric AML patients. These findings emphasise the importance of mtDNA copy number as a driver of disease biology. Among the 16 validated mitochondria-related DEGs analysed, we observed that the patients with higher mtDNA copy number had significantly higher expression of *SLC25A3, SDHC, RACK1,* and *FASTKD1* compared to patients with low mtDNA copy number. While, only a small percentage of mitochondrial proteins are coded by the mitochondrial genome, variations in mtDNA may modulate molecular signals through nuclear-mitochondrial crosstalk, which may promote tumorigenesis by upregulating oncogenes(34, 35). This suggests that in paediatric AML, cells with high mtDNA copy number are possibly driven through unique gene expression alterations, influencing disease biology and therapeutic response.

Comprehensive gene expression profiling has been extensively used to identify potential prognostic genes in adult AML; however, dysregulation of mitochondria-related gene expression, especially in children has not been well explored(36–38). Transcriptomic profiling of cytogenetically normal paediatric AML has identified complex genomic rearrangements and/or driver mutations in seemingly normal AML genomes and may even aid risk stratification(39, 40). Cai *et al*. developed a 3-gene prognostic risk model for children with AML using NCI TARGET dataset, although it was not externally validated(36). Similarly, Duployez *et al*. and Jiang *et al*. developed leukaemia stem cell score gene signature and immune checkpoint related gene signature respectively in paediatric AML predictive of survival outcomes(4, 41). None of the above studies evaluated alterations in mitochondrial gene expressions. Mitochondrial gene expression has been evaluated in other malignancies like ovarian cancer, where a mitochondria-related gene signature, consisting of 8 metabolic genes, has been identified with independent prognostic impact(42).

In this study, we identified exclusive mitochondria-related DEGs in paediatric AML and developed a prognostic gene signature including 3 genes (*SDHC, CLIC1,* and *SLC25A29).* The gene signature risk score was additionally found to be independently predictive of survival along with established ELN risk stratification with improved predictive ability over clinical risk categorization. The risk score was also found to be associated with poor clinical features of AML like the absence of RUNX1-RUNX1T1 translocation or poor-risk cytogenetics. Hence, the gene signature model is able to categorize the heterogenous molecular landscape of AML into clinically meaningful categories along with identification of adverse disease biology. The developed prognostic score also has the potential to identify high-risk subgroup even among those belonging to ELN good risk and vice-versa allowing better upfront risk stratification and personalized treatment decisions.

TCGA LAML dataset has been extensively used for identifying as well as validating prognostic gene signatures in various AML studies(43, 44). We used the LAML dataset of TCGA for external validation of our gene signature model and observed that the prognostic gene signature score was also independently predictive of survival outcome in a large adult cohort as well with predictive ability over and above known clinical predictors. This suggests that the identified DEGs have a prognostic impact in AML across age group.

Our gene signature included 3 mitochondria-related genes i.e., *SDHC*, *CLIC1*, and *SLC25A29*. *SDH* mutations lead to decreased activity of *SDH* with accumulation of succinate and increase in oxidative stress resulting in DNA damage and tumorigenesis(45). In contrast to previous findings, which suggests that the *SDH* gene is inactivated in solid tumors(46), we observed an increased expression of *SDHC* gene in AML which was predictive of worse survival. This is likely because, in contrast to solid malignancies, aggressive leukemias like AML depend on cellular oxidative phosphorylation for proliferation which is supported by upregulation of respiratory complex genes(47). Various studies also suggest dysregulation of chloride ion channels such as the *CLIC1* gene which plays a role in drug resistance and progression of various malignancies(22, 48). Although, the role of *CLIC1* in AML is still unexplored, we observed significant upregulation of *CLIC1* in paediatric AML with adverse prognostic impact. The downstream effects of upregulation of *CLIC1* on disease biology of AML need to be further deciphered.

In the current study, we observed an upregulation of *SLC25A29* in our cohort of paediatric AML patients, which is in line with previous studies where it was found to be significantly elevated in multiple malignancies(23). Similar upregulation was also been observed in adult AML patients of TCGA LAML dataset. However, on survival analysis, downregulation of *SLC25A29* was independently predictive of worse OS in our cohort. This finding was consistent even in the external cohort of TCGA LAML dataset, where even though the expression of *SLC25A29* was upregulated, a downregulated expression was predictive of worse survival outcomes. This finding was intriguing and the mechanism by which downregulation of *SLC25A29* drives a worse survival outcome remains unclear. *SLC25A29* is the main arginine transporter in the mitochondrial membrane(49). Aberrant upregulation of *SLC25A29* may result in transportation of more arginine into mitochondria, promoting synthesis of metabolites like nitric oxide, polyamines, proline and creatine, which are essential for cell survival and proliferation(50). Mitochondria-derived nitric oxide is known to have a dichotomous role in regulation of cancer progression which is influenced by expression of *SLC25A29*likelyaffecting disease outcome(51). The *SLC25* family of genes which encodes for a set of mitochondrial inner membrane carrier proteins, have been identified as a potential biomarker as well as novel therapeutic targets in various malignancies(52). The implications of altered expression of *SLC25A29* on disease biology of AML and its assessment as a therapeutic target is an exciting area of further research.

Our study has certain limitations. Transcriptomic profile and further validation by RTPCR were done in whole isolated mononuclear cell and not in sorted blasts. Initial selection of DEGs were also done from whole RNA sequencing of a limited number of samples, which may lead to a bias in selection, however, external validation of the validated genes confirmed their prognostic impact in an independent cohort.

In conclusion, this is the first study to report a validated set of mitochondria-related DEGs in paediatric AML. We observed that patients with high mtDNA copy number have a unique gene expression pattern possibly affecting disease biology. We developed a 3-gene based mitochondrial gene signature model with ability to predict prognosis in paediatric AML patients over and above established clinical prognostic parameters. The gene signature was also externally validated in a cohort of adult AML patients demonstrating its predictive ability in adult AML as well. Further directions for research include *in vitro* studies for elucidating the role of prognostic genes in leukemogenesis and their evaluation as potential targets for the treatment of paediatric AML.

## Data availability statement

The sequencing data is publicly available in repository (NCBI-SRA at PRJNA778747) and rest of data is available from the principal investigator on reasonable request.

## Data Availability

All data produced in the present study are available upon reasonable request to the author.

## Acknowledgements

The authors acknowledge the funding support from DST-SERB (Department of Science and Technology - Science and Engineering Research Board), Government of India for this work. (Extramural Research grant: EMR/2016/006376 and Core Research grant: CRG/2021/001887). The authors acknowledge the funding support from ICMR (Indian Council of Medical Research), Government of India for this work (ICMR SRF: 2019-6059/CMB/BMS). The authors also acknowledge every member of paediatric oncology team of our centre including research staff, nurses and dietician for their exemplary clinical services.

## Authorship Contribution Statement

Shilpi Chaudhary conceptualized the study, conducted the research, performed data analysis and interpreted results and wrote the manuscript. Shuvadeep Ganguly analysed data, interpreted results and wrote the manuscript. Jayanth Kumar Palanichamy, Archna Singh, Radhika Bakhshi and Anita Chopra conceptualized the study, provided intellectual inputs, administrative support and edited the manuscript. Dibyabhaba Pradhan conducted transcriptome data analysis. Sameer Bakhshi conceptualized the study, provided administrative support, intellectual inputs, interpreted results, wrote and edited the manuscript. All authors have reviewed and approved the final version of the manuscript.

